# Predicting age-related determinants of heterogeneous outcomes to COVID-19 mRNA vaccines through mathematical modelling

**DOI:** 10.1101/2025.02.14.25322308

**Authors:** Xiaoyan Deng, Suzan Farhang-Sardroodi, Morgan Craig

## Abstract

Older adults tend to exhibit weaker vaccine-elicited responses to mRNA COVID-19 immunization than younger people. This is a public health concern, as older individuals are more likely to experience severe COVID-19. To better understand the mechanisms of this age-related disparity, we developed a mathematical model of the post-vaccination humoral immune response. Through calibration to clinical data from 32 healthcare workers (HCWs) and 27 seniors who received the primary vaccine series (two priming doses and one booster), our model predicted that repeated vaccinations consistently enhanced antibody responses in both groups. While seniors were estimated to experience an accelerated decay in T follicular helper cells compared to HCWs, a larger booster dose effectively compensated for this weakened antibody response. Furthermore, we linked antibody and neutralization levels and used this relationship to predict post-vaccination neutralization, thus serving as a proxy for vaccine efficacy. By studying various combinations of mixed doses sizes in the primary vaccination series, our model predicted that administering a full-dose booster significantly enhances immunization outcomes, irrespective of the initial vaccine dose size. Further, a biannual half booster strategy was found to be more effective than one with an annual full booster, especially for seniors. Overall, our findings highlight the importance of tailoring vaccination strategies to different age groups to provide robust and long-lasting immunity against SARS-CoV-2 infections.

## INTRODUCTION

The fundamental goal of vaccination is to stimulate the immune system to establish long-term protection against severe illness and death upon pathogen re-exposure. Vaccine-induced B cell responses and the resulting production of neutralizing antibodies are crucial for preventing repeated infections by blocking viral entry and limiting viral replication. Since the emergence of COVID-19, multiple vaccines using different technologies have been approved against SARS-CoV-2 and its variants[1–5], with varying levels of efficacy and duration of protection. In this study, we focused on immune response dynamics following the primary three-dose mRNA (i.e., Comirnaty (Pfizer-BioNTech) and Spikevax (Moderna)) vaccine series with repeated boosters.

In clinical research, antibody titres are widely used as a measure of vaccine efficacy[6–8]. The potency and durability of this protection following immunization depends on multiple factors, such as race, sex, age, ethnicities, and individual health status[9–11]. Renia *et al.*[12] and Mwimanzi *et al.*[13] highlighted a notable difference in age-stratified antibody responses post-multiple COVID-19 vaccinations, finding that older people were more likely to have a relatively weaker, less effective and less durable vaccine-induced immunity following the first two mRNA COVID-19 vaccine doses[13]. However, both antibody responses and virus neutralization against the viral spike protein considerably improved after receiving a booster, reaching levels similar to those of younger healthcare workers (HCWs)[13]. The drivers of these distinct age-related antibody responses, the kinetics of booster vaccination on different aspects of the immune response, and the possible consequences of these boosters on the longevity of the ensuing immune response remain unclear.

To gain insight into age-related heterogeneity in vaccine responses between healthcare workers and seniors, we developed a mathematical model to investigate the mechanisms of vaccine-induced immunity and predict B cell memory responses after the primary mRNA COVID-19 vaccine series. Our model describes the initiation of humoral responses by vaccine particles, the generation of antigen-secreting cells including extrafollicular short-lived plasmablasts (PBs) and germinal center-dependent long-lived plasma cells (LLPCs), and the establishment and reactivation of B cell memory upon revaccination. We calibrated our model using *in vivo* and clinical data from a cohort of 32 HCWs and 27 seniors who received three COVID-19 mRNA vaccines in British Columbia, Canada[13]. Using our model, we identified factors responsible for the heterogeneous antibody responses between healthcare workers and seniors. Specifically, our results highlight an age-related disparity in T follicular helper (Tfh) cell kinetics between HCWs and seniors. Regardless of age, repeated vaccination was found to continuously enhance B cell activation and antibody production by plasmablasts and long-lived plasma cells. We also used our model to explore alternative vaccination strategies with varying dosing sizes to optimize age-specific protocols. We found that administering a full booster significantly improved neutralization outcomes following three-dose vaccination schedules. Further, our model predicted that implementing a biannual half-booster regimen after the primary series could maintain high neutralization protection in seniors compared to an annual full-booster schedule. This study therefore contributes to ongoing vaccination campaigns by rationalizing strategies to boost vaccine-induced humoral responses in key populations.

## METHODS

### Study cohort and data processing

In this study, we used data describing SARS-CoV-2 specific immune response from a cohort of COVID-19 vaccinees, including HCWs and seniors (aged 70 years and older) in British Columbia, Canada[13] to calibrate our mechanistic mathematical model of vaccine-induced responses (**Eqs. 1–10**). Specifically, the dataset included the measurements of total binding antibodies against SARS-CoV-2 spike (S) receptor binding domain in serum and plasma neutralization activity after Comirnaty (Pfizer-BioNTech) or Spikevax (Moderna). Serum and plasma were collected at seven specific time points: one month after the primary dose, and one, three, and six months after the second and the third doses, respectively (**Figure 1A**). Note that within this study cohort, all participants received two full doses during the initial two injections. However, seniors received the full dose at the third administration (booster), whereas healthcare workers were administered only half-doses[13].

**Figure 1.**
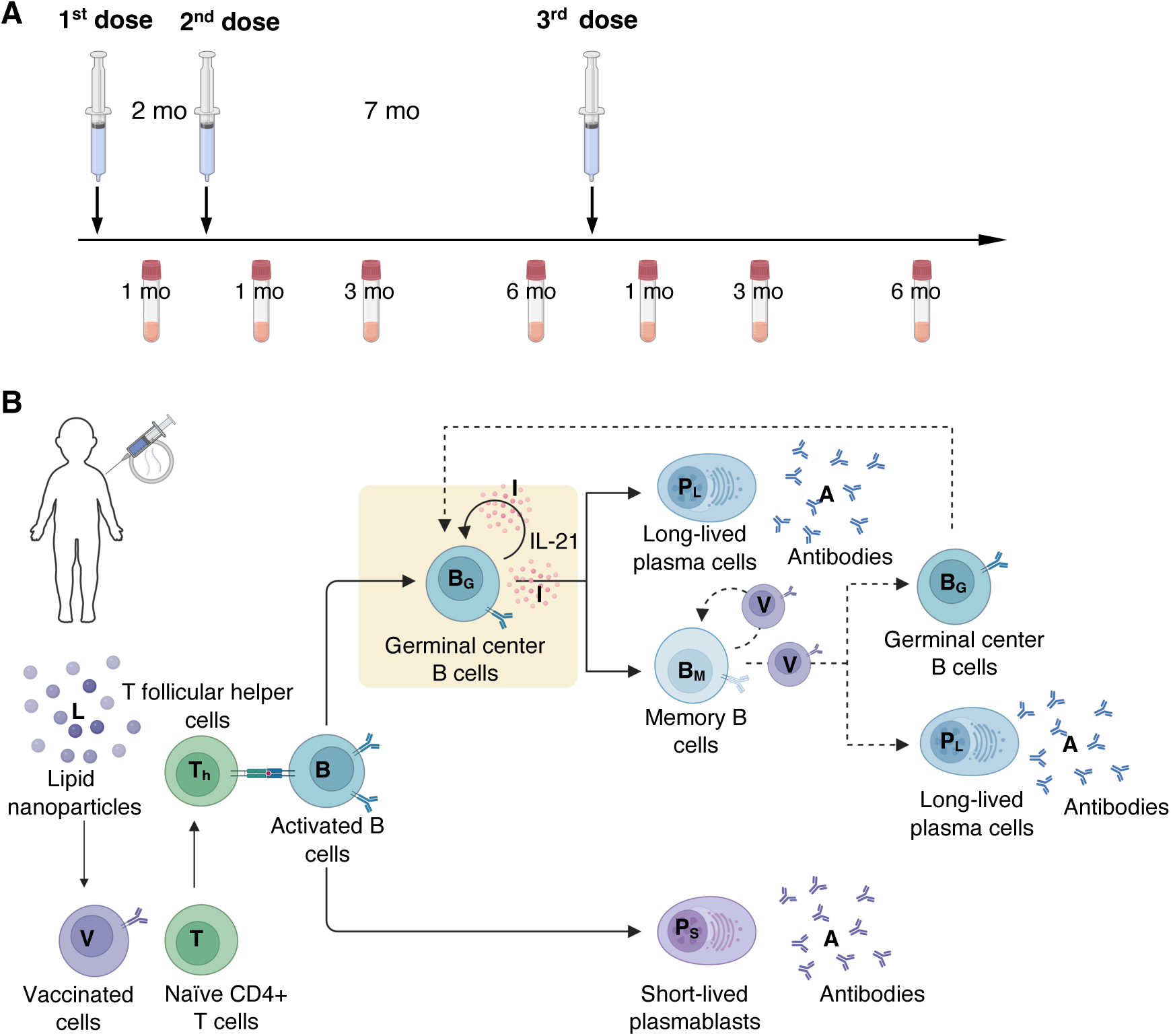
Illustrative representations of the vaccination protocol and mathematical model of the humoral response. **A)** Timeline of vaccine administration and sample collections. In the primary series considered in our study, the first two mRNA COVID-19 doses were administered at a fixed time interval of 2 months, with the third injected 7 months after the second[13]. Plasma was collected 1 month after the first dose and 1, 3, and 6 months after each of the following two doses. **B)** Schematic describing our mathematical model of the humoral immune response to vaccination. mRNA lipid nanoparticles (mRNA-LNPs) are initially taken up by nearby cells and delivered to target cells, transforming them into vaccinated cells (*V*) that promote naïve CD4+ T differentiation into T follicular helper cells (*T*_*h*_). These *T*_*h*_ cells, in turn, activate naïve B cells to become either plasmablasts (*P*_*S*_) or germinal center B cells (*B*_*G*_). Within the germinal center, B cells continue to develop into long-lived plasma cells (*P*_*L*_) and memory B cells (*M*), governed by IL-21 (*I*) secreted by T follicular helper cells. Both plasmablasts and LLPCs produce antibodies (*A*). Subsequent vaccine doses prompt memory B cells to either expand and differentiate or reinitiate the germinal center reaction in the presence of vaccinated cells. Solid arrows denote interactions that occur during all vaccine administrations, while dashed arrows represent processes that occur only with subsequent vaccinations.

Given that hybrid immunity enhances vaccine responses[14], we excluded any study participant with serological evidence of natural infection, as indicated by the presence of both anti-S and anti-N antibodies, to limit our study to individuals without prior exposure to the virus. Thus, our final cohort comprised individuals who only tested positive for anti-S antibodies. Moreover, we excluded one patient with only two neutralization data points and another with extremely high antibody concentrations but low neutralization levels. Consequently, our study included a total of 59 participants (32 HCWs and 27 seniors).

For model calibration, we ignored nominal time and assumed a uniform fixed dose interval of one month after the first dose and seven months after the second dose for all individuals (**Figure 1A**).

In cases where multiple neutralization data points were available for the same visit, we calculated their average values for analytical precision. Moreover, we excluded neutralization data from the fourth visit if it was higher than the values from the third visit, as most individuals showed a decline, making such increases biologically implausible.

### Mathematical model of the humoral immune response to COVID-19 vaccination

To quantitatively study the humoral memory response after primary series mRNA COVID-19 vaccination, we developed a mechanistic mathematical model (**Eqs. 1–10**) that describes how mRNA vaccine particles stimulate the humoral immune response, establish antigen-specific immunological memory, and affect immune responses after subsequent vaccine administrations (**Figure 1B**).

Upon initial intramuscular injection, mRNA molecules encapsulated in lipid nanoparticles (*L*) are absorbed by muscle cells near the injection site and transported via blood vessels to target cells. Once at their target sites, the mRNA molecules are released from the lipid nanoparticles and translated into the SARS-CoV-2 spike protein, converting host cells into vaccinated cells (*V*) at a rate of *δ*_*LV*_. These vaccinated cells are then recognized by the immune system, initiating the differentiation of naïve CD4+ T cells into T follicular helper cells (*T*_*h*_) at a rate of *δ*_*TV*_ [15]. Subsequently, naïve B cells, activated by Tfh cells at rate *δ*_*BT*_, either migrate into germinal centers (GC), forming germinal center B cells (*B*_*G*_) at rate *ρ*_*G*_, or evolve directly into GC-independent plasmablasts (*P*_*S*_) at per capita growth rate *ρ*_*S*_[16–18]. Interleukin-21 (IL-21; *I*), secreted by *T*_*h*_cells at rate *ρ*_*I*_, regulates the proliferation and differentiation of GC B cells[19–23] in a saturating, sigmoidal effect 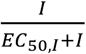 with a maximum rate *δ*_*IG*_. Correspondingly, GC B cells proliferate at their maximal rate *β*_*G*_ and undergo either self-renewal or differentiation with probabilities *p*_*B*_ and (1 − *p*_*B*_), respectively. Upon differentiation, they further develop into antigen-specific long-lived plasma cells (*P*_*L*_) or memory B cells (*M*) with probability *p*_*P*1_or (1 − *p*_*P*1_), respectively. Plasma B-cells are long-lived, non-proliferating cells arising from B-cell differentiation, stimulated by interaction with T follicular helper cells. Both plasmablasts and long-lived plasma cells secrete antibodies (*A*) at production rates *α*_*S*_ and *α*_*P*_, respectively[24].

Upon subsequent vaccination, vaccinated cells trigger pre-existing memory B cells to quickly proliferate at rate *β*_*M*_. During proliferation, these cells either self-renew or differentiate with probabilities *p*_*M*_ or (1 − *p*_*M*_), respectively. During the differentiation phase, they may either progress into long-lived plasma cell with probability *p*_*P*2_ or revert to germinal center B cells with probability (1 − *p*_*P*2_), thereby triggering the secondary germinal center reaction[25]. All cells, cytokines, and particles decay naturally at rates *d*_*X*_, where *X* is the corresponding variable.

The complete model is described by

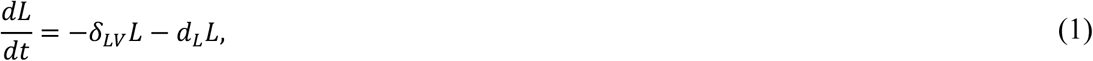

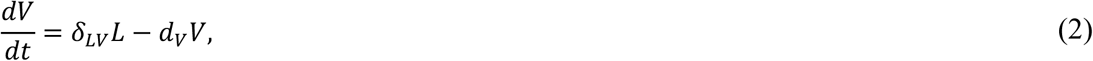

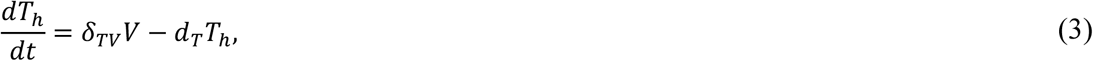

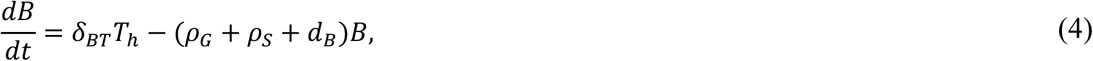

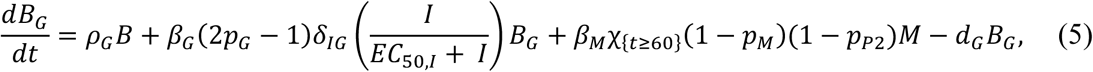

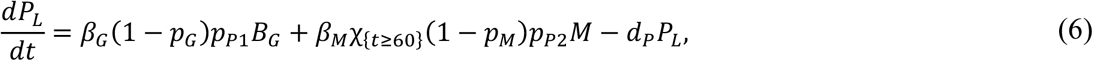

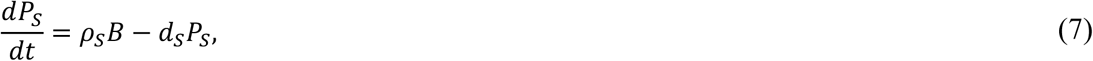

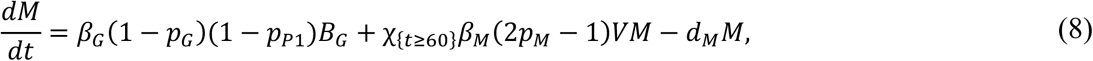

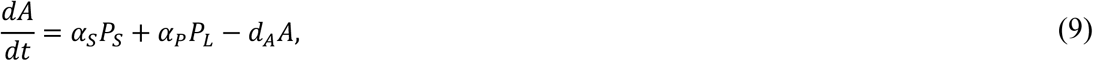

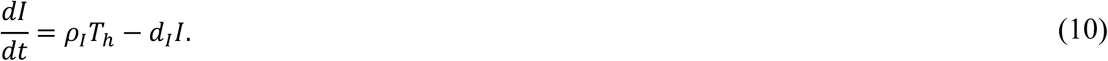

The indicator functions in Equations 5 and 8, i.e.,

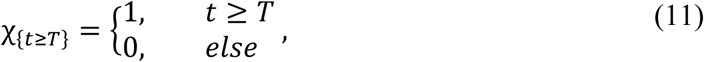

describe immunological boosting upon repeated vaccine administrations at time *T*.

### Uncertainty and sensitivity analysis

To identify parameters with significant nonlinear yet monotonic correlations to antibody dynamics after multiple vaccinations, we performed a global sensitivity analysis using Partial Rank Correlation Coefficients (PRCC)[26] and explored the impact of parameter variability on antibody responses after the second vaccine dose. Based on average parameter estimates, we generated 1000 distinct parameter by varying each parameter between half and twice its average value using Latin hypercube sampling. We then calculated PRCC values and measured the relationship between each model parameter and the maximum antibody concentrations after administration of primary two doses.

### Estimation of heterogeneous antibody responses after multiple vaccinations

Model parameters were estimated using data from the literature and antibody and neutralization data from Mwinmazi *et al*.[13] (**Supplementary section: Parameter estimation**). Given that there were no available pharmacokinetic data, we kept the vaccine pharmacokinetics consistent across all individuals and doses.

To characterize individual antibody waning patterns, we varied the Tfh cell decay rate (*d*_*T*_), as the global sensitivity analysis identified it as the second most influential factor negatively affecting antibody peaks after the initial doses (**Figure 1A**). We used *λ*_1_, *λ*_2_, and *λ*_3_ to increase the parameters found to be positively correlated with peak antibody concentrations to mimic antibody concentration boosting following each of three doses. Specifically, *λ*_1_ was set to 1, representing the initial immune response primed by the first dose, while *λ*_2_ and *λ*_3_were determined using antibody data collected after the second and third doses, respectively. We then incorporated these amplification factors through the indicator functions *χ*_{*t*≥*T*}_ (Eq. 11) and the term B*λ*_1_ + *χ*_{*t*≥60}_*λ*_2_ + *χ*_{*t*≥270}_*λ*_3_D in Eqs. 12-13. The latter represents the cumulative immunological amplification after the second and third doses administered on days 60 and 270, respectively. Kocher *et al*.[27] reported that a hypervaccinated individual who received 215 vaccines exhibited higher peak antibody titers that eventually became comparable to those of standard vaccinees, with only a modest increase after two additional vaccinations, suggesting a potential immunological ‘ceiling’. Thus, we assumed that immunological amplification of the entire population remains comparable after each subsequent dose. In addition, although the production rate of Tfh cells induced by vaccination (*δ*_*TV*_) was strongly correlated with antibody peaks, we fixed this value, given that multiple vaccine doses do not significantly boost T cell responses[28,29]. Taken together, we selected three specific parameters for amplification: *δ*_*BT*_ (generation rate of activated B cells by Tfh cells), *α*_*S*_ (antibody production rate by plasmablasts), and *α*_*p*_ (antibody production rate by long-lived plasma cells), and extended our model by boosting the activation of B cells in Eq. 4 and two sources of antibody production Eq. 9 as follows:

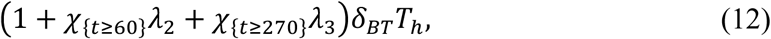

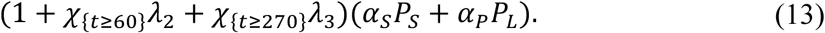

To capture vaccine response dynamics for each individual in both age groups, we fixed all other parameter values and estimated the Tfh decay rates (*d*_*T*_) along with two amplification factors (*λ*_2_and *λ*_3_ in Eqs. 12-13). These three parameter were estimated a non-linear mixed-effects model in Monolix 2024 R1, as in previous studies[30–32]. Importantly, the model fit was found to have a lower Bayesian information criterion value compared to models that did include amplification of these three rates (*δ*_*BT*_, *α*_*S*_, and *α*_*p*_) (**Supplementary Table 2**).

### Estimation of viral neutralization as a function of anti-S antibody concentrations

Virus neutralization is directly linked to circulating antibody concentrations[33–36], so we quantified their relationship using the usual sigmoidal stimulatory model[37,38] given by

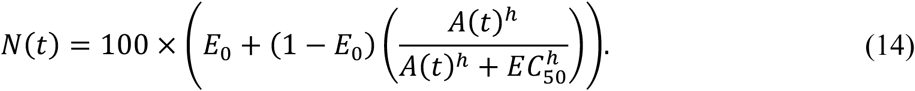

Here, *N*(*t*) is neutralization over time, *E*_0_ represents the baseline neutralization before receiving second dose, *A*(*t*) is the time-dependent antibody level, *h* is the Hill coefficient that measures the steepness of the neutralization curve, and *EC*_50_ denotes the antibody concentration inducing 50% neutralization between the baseline and maximum levels (here, the maximum neutralization was assumed to be 100%).

To estimate the values of *E*_0_, *EC*_50_ and *h* in Eq. 14 for each individual, we used average and individual antibody and neutralization data Mwimanzi *et al.* using a non-linear mixed-effects model in Monolix.

### Statistical analysis

The Pearson correlation coefficient was used to measure the strength and direction of the linear relationship between two variables, implemented using the *corrcoef* function in MATLAB R2022a. Correlations were considered statistically significant at the α =5% significance level. To compare two distributions, we used the two-tailed Wilcoxon-Mann-Whitney test and the two-tailed Kolmogorov-Smirnov test. The Wilcoxon-Mann-Whitney test, performed via the *Wilcox.Test* function in RStudio 2024.9.0.375, is a nonparametric alternative to the two-sample t-test without assuming normality. The Kolmogorov-Smirnov test, implemented using the *kstest2* function in MATLAB R2022a, is used to detect differences in the shape or dispersion of distributions. Both tests were performed with a level of significance of α =5%.

## RESULTS

### Dose size-dependent comparable vaccine-induced immune responses after the booster in seniors and healthcare workers

Using our mechanistic mathematical model of vaccine-induced responses (**Eqs. 1–10**), we first aimed to understand dose size-independent differences in humoral responses between healthcare workers and seniors. For this, we performed a sensitivity analysis (see **METHODS**) to examine how model parameters influence key antibody metrics (i.e., peak concentrations after each of the first two mRNA vaccine doses). This analysis showed the activation rate of B cells by Tfh cells (*δ*_*BT*_) and the production rate of Tfh cell stimulated by vaccinated cells (*δ*_*TV*_) to be strongly positively correlated with peak antibody concentrations after the two priming doses, with partial rank correlation coefficients (PRCC) exceeding 0.8 after the first and 0.7 after the second dose, respectively (*p* < 0.05). The PRCC value of antibody production rates by plasmablasts (*α*_*S*_) dropped from 0.79 to 0.58 from the first to second dose. Conversely, the antibody secretion rate by long-lived plasma cells (*α*_*P*_), a key driver of sustained antibody production, showed a weak correlation with the first antibody peak (PRCC = 0.22) but a slightly stronger correlation with the second peak (PRCC = 0.30). Additionally, decay rates of vaccinated cells (*d*_*V*_) and Tfh cells (*d*_*T*_) had significant negative associations with peak antibody values for first two doses (PRCCs < -0.7, *p* < 0.05) (**Figure 2A and Supplementary Figure 1A–B**). PRCC scatter plots confirmed both the positive and negative correlations of these parameters with respect to the first and second antibody peaks (**Supplementary Figure 1C–D**).

**Figure 2.**
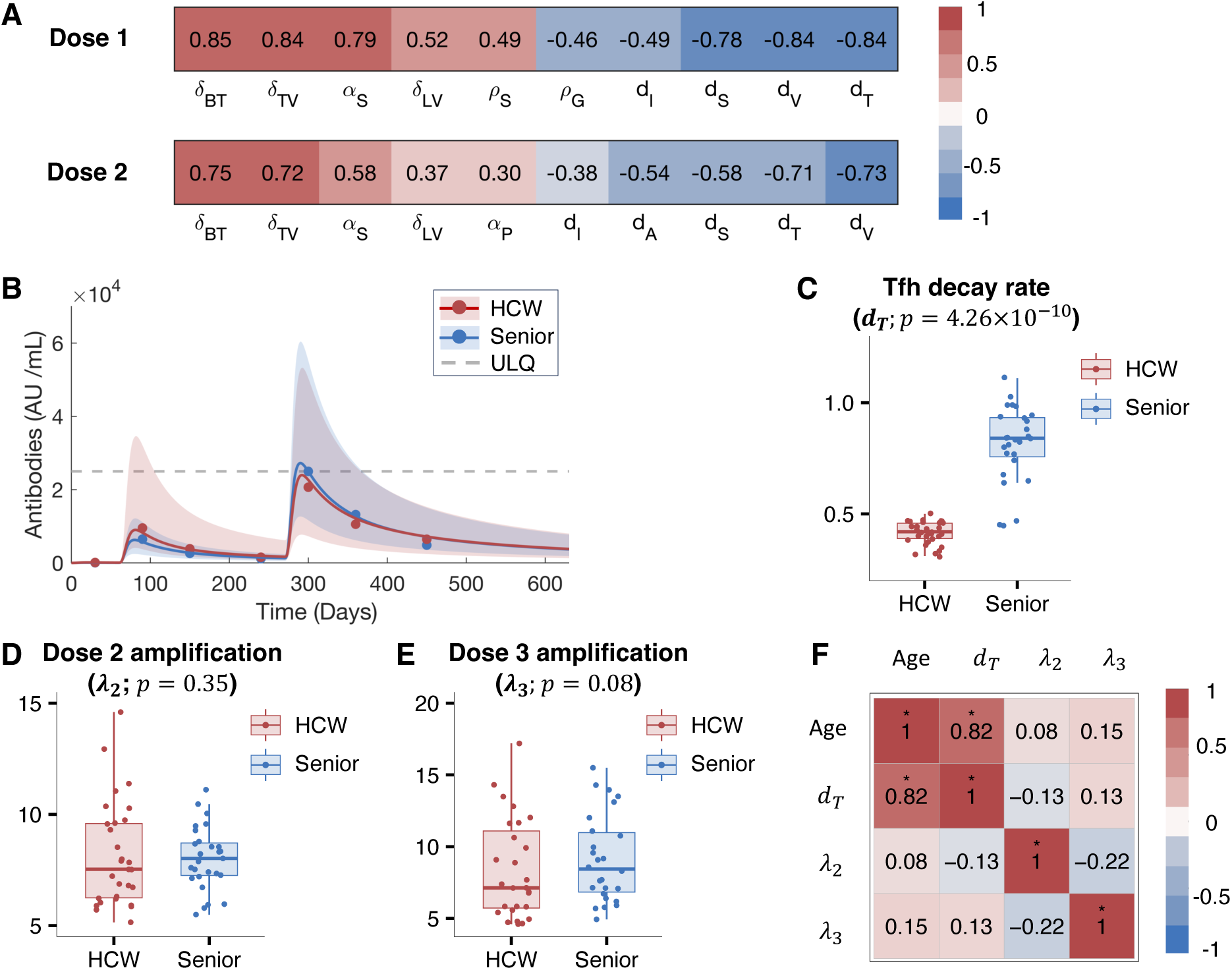
Age is positively correlated with the decay rate of T follicular helper cells. **A)** Global sensitivity analysis results reported as partial rank correlation coefficients with respect to peak antibody concentrations after the first and second dose. Shown are the ten most statistically significant parameters (*p* < 0.05). **B)** Estimated antibody dynamics between HCWs and senior. Red: health care workers. Blue: seniors. Filled circles: median of observed antibody data from Mwimanzi *et al.*[13]. Solid lines: median model-predicted antibody concentrations. Dashed gray line: upper limit of quantification. Shaded region: 95% confidence interval of predicted antibody dynamics. **C-E)** Boxplots of Tfh cell decay rates ( *d*_*T*_), vaccine-induced immunological amplification after the second dose (*λ*_2_), and immunological boost after dose 3 (*λ*_3_) for HCWs and seniors, respectively. Filled circles: individual predicted values. Indicated p-values calculated using a Wilcoxon-Mann-Whitney test. **F)** Correlation matrix between age and the three estimated parameter values (*d*_*T*_, *λ*_2_ and *λ*_3_). Pearson correlation values are indicated by the size and value of the squares, and * indicates the 5% significance level.

Based on these findings, we then individualized our model using antibody data from both the HCW and senior cohorts described in Mwimanzi et al.[13] (see **METHODS** and **Supplementary Figure 2A–B**). Compared to the senior cohort, healthcare workers on a whole were predicted to exhibit higher and more variable median antibody concentrations following two priming doses, with lower peak median concentrations post-boost (median peak levels: 2.40 × 10^4^ AU/mL in HCWs vs. 2.72 × 10^4^AU/mL in seniors; **Figure 2B**). It is important to note that in our study’s cohort[13], HCWs received a half-dose booster while seniors were administered a full dose, per national vaccination guidelines. We posited that this difference in dose size could explain the higher and more sustained antibody titres observed in seniors compared to younger individuals. To confirm this, we used our model to simulate both groups receiving full booster doses. As expected, this resulted in more robust vaccine responses in healthcare workers. Indeed, the median antibody peak level in HCWs was predicted to be about 1.6 times higher than in seniors ( 4.36 × 10^4^AU/mL vs. 2.72 × 10^4^AU/mL). Additionally, HCWs were predicted to have elevated levels of two key components of long-term immunity—memory B cells and long-lived plasma cells—with median levels approaching the upper 95% confidence interval of the senior cohort (**Supplementary Figure 2B–G**).

### Accelerated T follicular helper cell decay observed in seniors versus healthcare workers

Based on our global sensitivity analysis, the Tfh cell decay rate (*d*_*T*_), and the immunological amplification factors after the second and third doses, (*λ*_2_ and *λ*_3_, respectively) could effectively capture differential antibody response patterns across the entire population (**Supplementary Figure 2A–B**). Comparing individual estimated *d*_*T*_values between HCWs and seniors, we observed a statistically significant difference of *p* = 4.26 × 10^−10^ (Wilcoxon-Mann-Whitney test, significance level *α* =5%) between the two age groups. Specifically, seniors were predicted to have faster decay rates, with median values double those of HCWs (0.84 vs. 0.42 per day) and significantly greater variability, with a standard deviation about three times larger (0.17 vs. 0.05) (**Table 1**). Moreover, we identified a strong positive relationship between age and the Tfh cell decay rate, with a Pearson correlation coefficient of 0.82 and a *p*-value of 2.27 × 10^−15^ (**Figure 2C**).

**Table 1.**
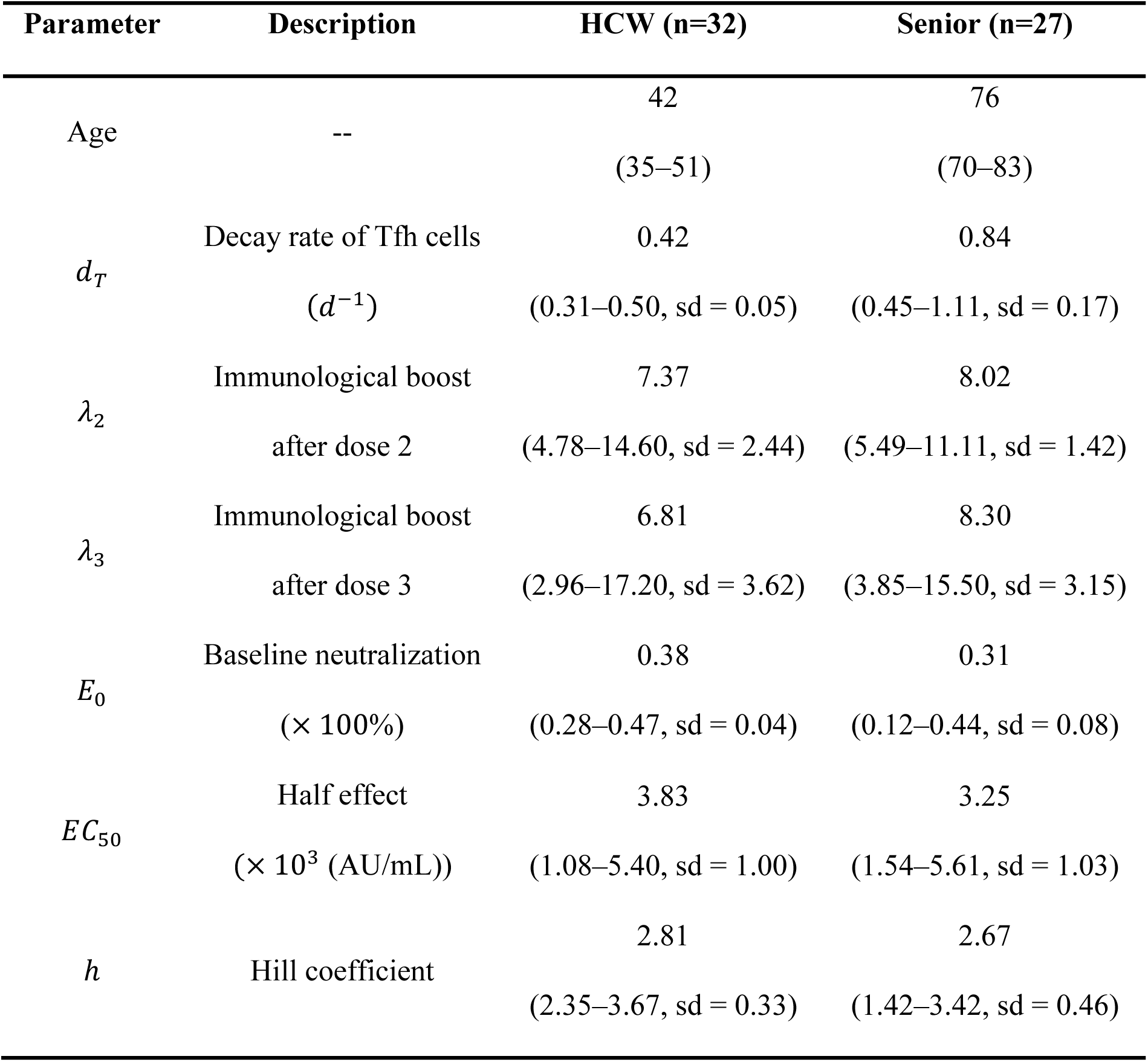
Estimated cohort and individual parameters values. Th decay rates and amplification factors were fit using individual antibody concentrations for HCWs and seniors, with all other parameter values fixed. Parameter values of antibody-neutralization relationship (Eq. 14), *E*_0_, *h*, and *EC*_50_, were fit to individual antibody titres and virus neutralization in seniors and healthcare workers. Median estimates are indicated with ranges and standard deviations (sd).

| Parameter | Description | HCW (n=32) | Senior (n=27) |
| --- | --- | --- | --- |
| Age | -- | 42<br>(35–51) | 76<br>(70–83) |
| $d_T$ | Decay rate of Tfh cells<br>($d^{-1}$) | 0.42<br>(0.31–0.50, sd = 0.05) | 0.84<br>(0.45–1.11, sd = 0.17) |
| $\lambda_2$ | Immunological boost<br>after dose 2 | 7.37<br>(4.78–14.60, sd = 2.44) | 8.02<br>(5.49–11.11, sd = 1.42) |
| $\lambda_3$ | Immunological boost<br>after dose 3 | 6.81<br>(2.96–17.20, sd = 3.62) | 8.30<br>(3.85–15.50, sd = 3.15) |
| $E_0$ | Baseline neutralization<br>($\times 100\%$) | 0.38<br>(0.28–0.47, sd = 0.04) | 0.31<br>(0.12–0.44, sd = 0.08) |
| $EC_{50}$ | Half effect<br>($\times 10^3$ (AU/mL)) | 3.83<br>(1.08–5.40, sd = 1.00) | 3.25<br>(1.54–5.61, sd = 1.03) |
| $h$ | Hill coefficient | 2.81<br>(2.35–3.67, sd = 0.33) | 2.67<br>(1.42–3.42, sd = 0.46) |

Our analysis further revealed no statistically significant differences in the amplification parameters *λ*_2_ and *λ*_3_ between seniors and healthcare workers, nor any significant Pearson correlation between these values and age (**Figure 2D–F**). However, HCWs demonstrated greater variability in immunological boosts after multiple doses compared to seniors (**Supplementary Figure 2I** and **Table 1**). Notably, vaccine-induced antibody responses were predicted to be continuously enhanced from the primary dose through the booster. Specifically, starting from the baseline value of 1, the median cumulative fold increases rose comparably to 8.37-fold in HCWs and 9.02-fold in seniors after the second dose and further increased to statistically similar levels of 16.36-fold and 17.15-fold after the booster, respectively (Wilcoxon-Mann-Whitney test, significance level *α* =5%, *p* = 0.15; **Table 1** and **Supplementary Figure 2I–J**). These findings highlight that repeated vaccination progressively strengthened immunity, providing a robust and age-independent immunological boost for all participants.

### Enhanced neutralization durability and reduced group disparity after booster

To evaluate vaccine-induced protection, we next correlated sampled antibody concentrations to neutralization levels from Mwimanzi et al[13] via our established antibody-neutralization relationship (**Eq. 14)**. We found that neutralization at baseline *E*_0_(i.e., neutralization before receiving the second dose) varied significantly between HCWs and seniors, with seniors displaying a significant lower median value (31% vs. 38%; *p* = 2.21 × 10^−5^; **Supplementary Figure 3A–B**). Additionally, this baseline value was negatively correlated with age (Pearson correlation of −0.52 and *p* = 2.08 × 10^−5^; **Table1** and **Supplementary Figure 3C and F**), indicating an overall weaker immune response in seniors after the first priming dose. Conversely, as expected, no statistically significant correlations were observed between age and the estimates of *EC*_50_ (half effect) and *h* (Hill coefficient), nor were there significant differences in these parameters between the two age groups (**Supplementary Figure 3D–F**).

Using each participant’s predicted antibody dynamics, we then generated personalized neutralization trajectories (**Supplementary Figure 3G–H**). On the whole, repeated vaccination was found to significantly improve neutralizing responses, achieving higher and more sustained peak levels with slower declines. Compared to the senior cohort, HCWs exhibited greater variability in after-peak neutralization levels (**Figure 3A** and **Supplementary Figure 3G–H**). Median neutralization levels steadily increased from 38% in HCWs and 31% in seniors before the second dose to 46.43% and 36.75% just before the booster. One year after the booster, median neutralization further rose to 67.72% in HCWs and 65.54% in seniors **(Figure 3B–D)**. Though the distributions of neutralization levels for each age group were statistically different after the first and second doses (*p* < 0.05), this difference disappeared after the booster (*p* = 0.98) based on a Kolmogorov-Smirnov test **(Figure 3B–D)**.

**Figure 3.**
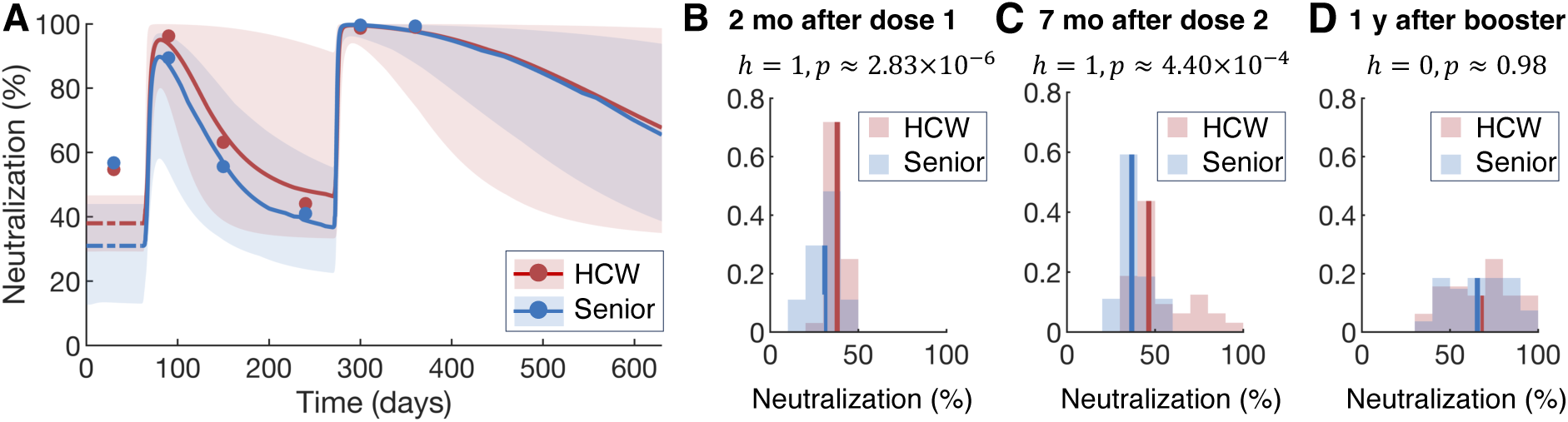
Predicted neutralization dynamics and distribution of neutralization levels for HCWs and seniors. **A)** Model predicted neutralization dynamics in HCWs and seniors. Filled circles: median of sampled neutralization data from Mwimanzi et al[13]. Solid lines: median predicted neutralization. Shaded regions: 95% confidence intervals of model-predicted neutralization. **B)** Distribution of neutralization levels 2 months after the primary dose, prior to the second dose. **C)** Distribution of neutralization levels 7 months after second dos, prior to booster. **D)** Distribution of neutralization one year after the booster. **B–D)** Values of *h* and *p* were calculated using the Kolmogorov-Smirnov test. Vertical lines: median neutralization. Blue: seniors. Red: healthcare workers.

To further examine differences in vaccine-induced protection between groups, we evaluated the duration of neutralization remained above chosen thresholds of 60%, 70%, 80%, 90%, and 95%. We found that the booster significantly extended this duration in both age groups, even when HCWs received only a half booster dose. Both HCWs and seniors were able to reach the high threshold levels of 80%, 90%, and 95%, with nearly all vaccinated individuals—except for one HCW—achieving 95% neutralization for a mean duration of over 4 months post-booster (**Supplementary Figure 4**). In contrast, 36 study participants could not achieve this neutralization threshold after the first two doses, underlining the importance of the three-dose primary series. Further, 21 HCWs and 18 seniors (representing around 66% of the entire study cohort) sustained neutralization above 80% for over 200 days, while more than half maintained levels above 90% for over half a year after the booster. For the 60% and 70% thresholds, the median duration increased significantly from approximately 2–3 months after the second dose to over 330 days post-booster for the entire population (**Table 2**). Notably, while the booster improved the immunological profiles of seniors, none were able to sustain the 95% neutralization threshold throughout the year, even after receiving the full booster, which suggests that an additional dose may be required within one year if the threshold is set to 95% (**Supplementary Figure 4B**).

**Table 2.** Predicted duration above neutralization thresholds post-second and third doses. Calculated median duration above 60%, 70%, 80%, 90%, and 95% after the 2nd and 3rd doses. The range in each cohort is provided in the parentheses.

| Neutralization<br>threshold | Duration above the threshold<br>after dose 2 (days) |  | Duration above the threshold<br>after the booster (days) |  |
| --- | --- | --- | --- | --- |
|  | HCWs | Seniors | HCWs | Seniors |
| 60% | 107.5 (44–210) | 83 (0–179) | 360 (126–360) | 360 (222–360) |
| 70% | 81 (34–210) | 62 (0–125) | 342.5 (101–360) | 330 (173–360) |
| 80% | 62 (0–210) | 45 (0–89) | 274 (72–360) | 263 (126–360) |
| 90% | 41 (0–210) | 0 (0–59) | 191.5 (40–360) | 186 (76–360) |
| 95% | 13.5 (0–186) | 0 (0–39) | 134 (0–360) | 128 (35–334) |

### Significant influence of booster dose size on sustained neutralization durability following primary vaccination series

Our previous analyses showed that a full booster enabled seniors to achieve neutralization levels comparable to HCWs who received a half booster. Building on these findings, we aimed to further explore how dose size determines vaccine outcomes of the three-dose primary series. Specifically, we simulated eight combinations of mixed half-dose (‘H’) and full-dose (‘F’) using the same dosing intervals as Mwimanzi *et. al*[13] (**Figure 4A**) and predicted the resulting neutralization for one year post-primary series. As expected, HCWs exhibited more robust neutralization responses than seniors under the same vaccination schedule, achieving higher and longer-maintained neutralization (**Figure 4B**). For instance, HCWs were predicted to sustain median neutralization above 90% for about 290 days—roughly 102 days longer than seniors—after three full doses (‘FFF’); median neutralization levels were similarly higher (84.14% vs. 65.54%) in HCWs one-year post primary series. Interestingly, the initial dose size was predicted to have minimal impact on long-term neutralization, as shown by the overlapping neutralization curves for regimens with different initial doses but identical subsequent doses (**Supplementary Figure 5**). However, the size of the booster dose was found to significantly influence outcomes. For example, regimens with a full booster (‘FHF’ and ‘FFF’) were predicted to consistently achieve higher median neutralization levels than those with half dose boosters (‘FHH’ and ‘FFH’). Similarly, the ‘FHF’ regimen outperformed ‘FFH’. These results further highlight the critical role of a third full dose booster for infection protection in both HCWs and seniors (**Figure 4B**).

**Figure 4.**
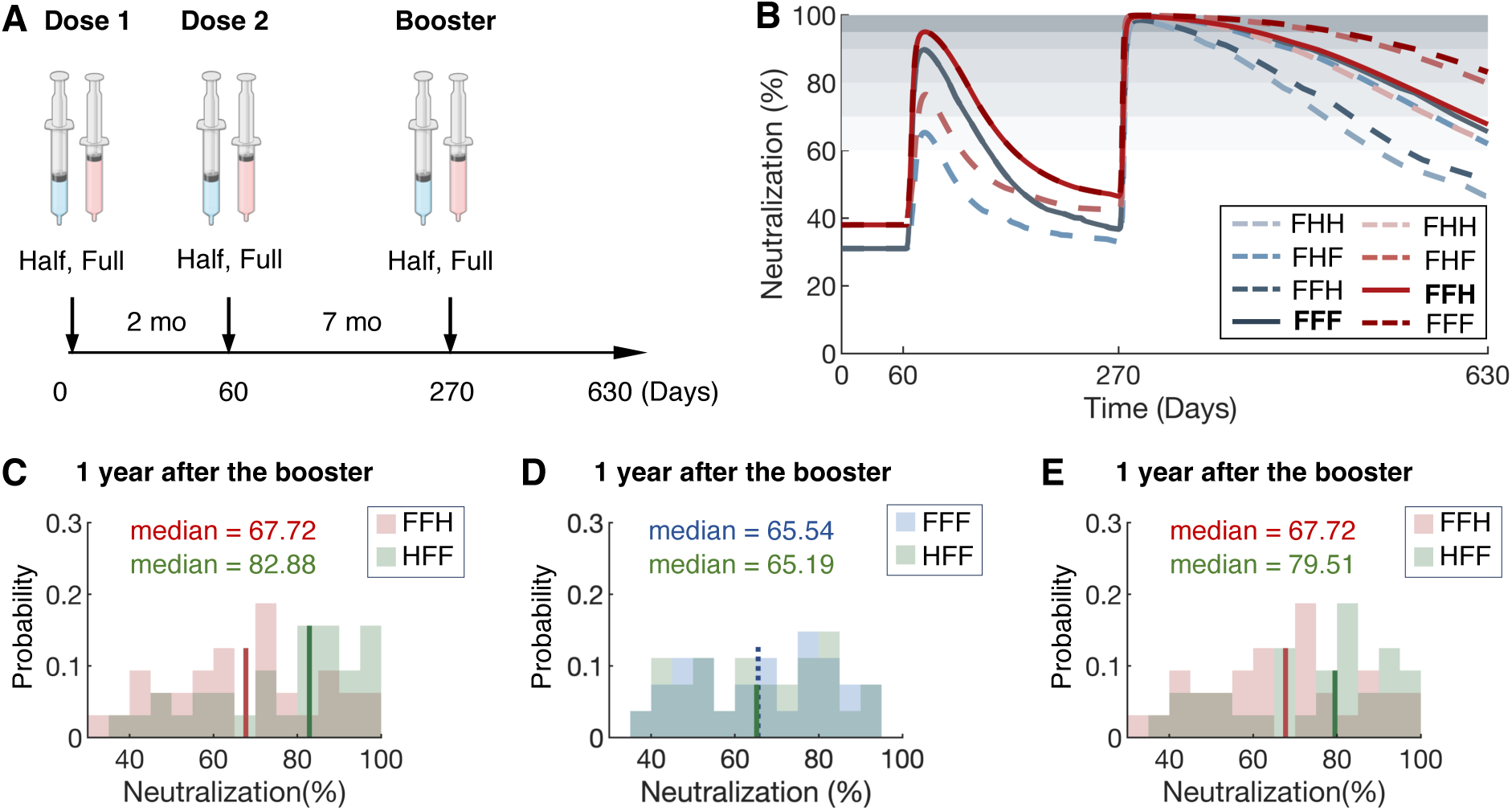
Dosing strategies impact neutralization dynamics in health care workers and seniors. **A)** Schematic describing approach to study mixed vaccination schedules with either full (pink; F) and half (blue; H) doses, with a 2-month interval between the first two doses, and 7 months between the second dose and booster (Mwimanzi et al.[13]). **B)** Predicted median neutralization curves for HCWs and seniors based on four mixed-dose regimens: ‘FFF’ (three full doses), ‘FFH’ (two full priming doses followed by a half-dose booster), ‘FHF’ (the full first and third doses with a half-dose second), and ‘FHH’ (a full dose followed by two half-doses). Red: health care workers. Blue: seniors. Solid lines: neutralization after primary series per the guidelines described in Mwimanzi *et al*.[13]. Dashed lines: model predicted neutralization outcomes. Shaded gray regions: neutralization thresholds of 60%, 70%, 80%, 90%, and 95%. **C)** Distribution of neutralization levels one year after three doses under the ‘FFH’ and ‘HFF’ regimens in HCWs. **D)** Distribution of neutralization levels one year after three doses under the ‘FFH’ and ‘HFF’ regimens in seniors. **E)** Distribution of neutralization levels one year after three doses under the ‘FFH’ and ‘HHF’ regimens in HCWs. **C-E)** Vertical lines and dashed lines: median values.

Indeed, our model suggested that the ‘HFF’ strategy (a half dose followed by two full boosters) more effectively enhanced neutralization responses compared to the ‘FFH’ (two full doses followed by a half booster) schedule in HCWs by prolonging neutralization levels above all thresholds, increasing the number of fully immunized individuals, defined as those achieving neutralization above each threshold throughout the dosing interval, and raising the median neutralization in the HCW cohort from 67.72% to 82.88% one year after the third dose (**Figure 4C** and **Supplementary Figure 6A**). Specifically, following the ‘HFF’ vaccination series, HCWs experienced an increase of over 3 months in the median time above the 80% and 90% neutralization thresholds (from 274 to 360 days and 191.5 to 286 days, respectively), as well as an additional 76.5 days above the 95% threshold. Moreover, the proportion of fully immunized individuals at least doubled for the 80%, 90%, and 95% thresholds. (**Supplementary Figure 6A**). Similarly, the ‘HFF’ regimen provided seniors with protection comparable to the ‘FFF’ guideline, with comparable neutralization durability and nearly identical neutralization levels one-year post-booster (65.19 % vs. 65.54%; **Figure 4D** and **Supplementary Figure 6B**). Among HCWs, the ‘HHF’ vaccination series (two half priming doses followed by a full booster) also improved outcomes, prolonging protection above thresholds and increasing the number of fully immunized individuals, though it was less effective than the ‘HHH’ regimen (**Supplementary Figures 6C–D**). Notably, both ‘HHF’ and ‘HFF’ regimens achieved comparable high median neutralization levels (79.51% vs 82.88%) in the HCW cohort one-year post-booster.

### High neutralization maintained with annual full boosters or biannual half-dose boosters

We calibrated our model to a clinical study[13] focused on the three-dose primary mRNA COVID-19 vaccination series (2 priming doses + 1 booster) performed between December 2020 and January 2022. Since then, SARS-CoV-2 has become endemic and public health agencies have varying recommendations on booster schedules for seniors, individuals with comorbidities, healthcare workers, and healthy younger individuals[39–41]. Therefore, we sought to use our model to determine the most effective strategy for maintaining high viral neutralization levels through repeated vaccination. According to Kocher *et al*., an individual who received 217 COVID-19 vaccines[27] maintained functional quality of adaptive immune responses comparable to that of standard vaccinees without displaying constant amplification in their responses. Therefore, we assumed no increase in immunological amplification after the primary series of three vaccines. We next used our model to predict neutralization responses under two scenarios: (1) younger individuals (HCWs) receiving a half-dose booster and seniors receiving a full-dose booster annually (similar to the primary series dosing strategy in Mwimanzi *et al*.[13]), and (2) both age groups receiving either half- or full-dose boosters annually. Additionally, we compared these scenarios to an alternative strategy for seniors involving annual full-dose boosters combined with biannual half-dose boosters, starting one year after the initial booster (dose 3). Neutralization levels were quantified yearly—either one year after the annual booster or six months after every two biannual half-dose boosters (**Figure 5A**).

**Figure 5.**
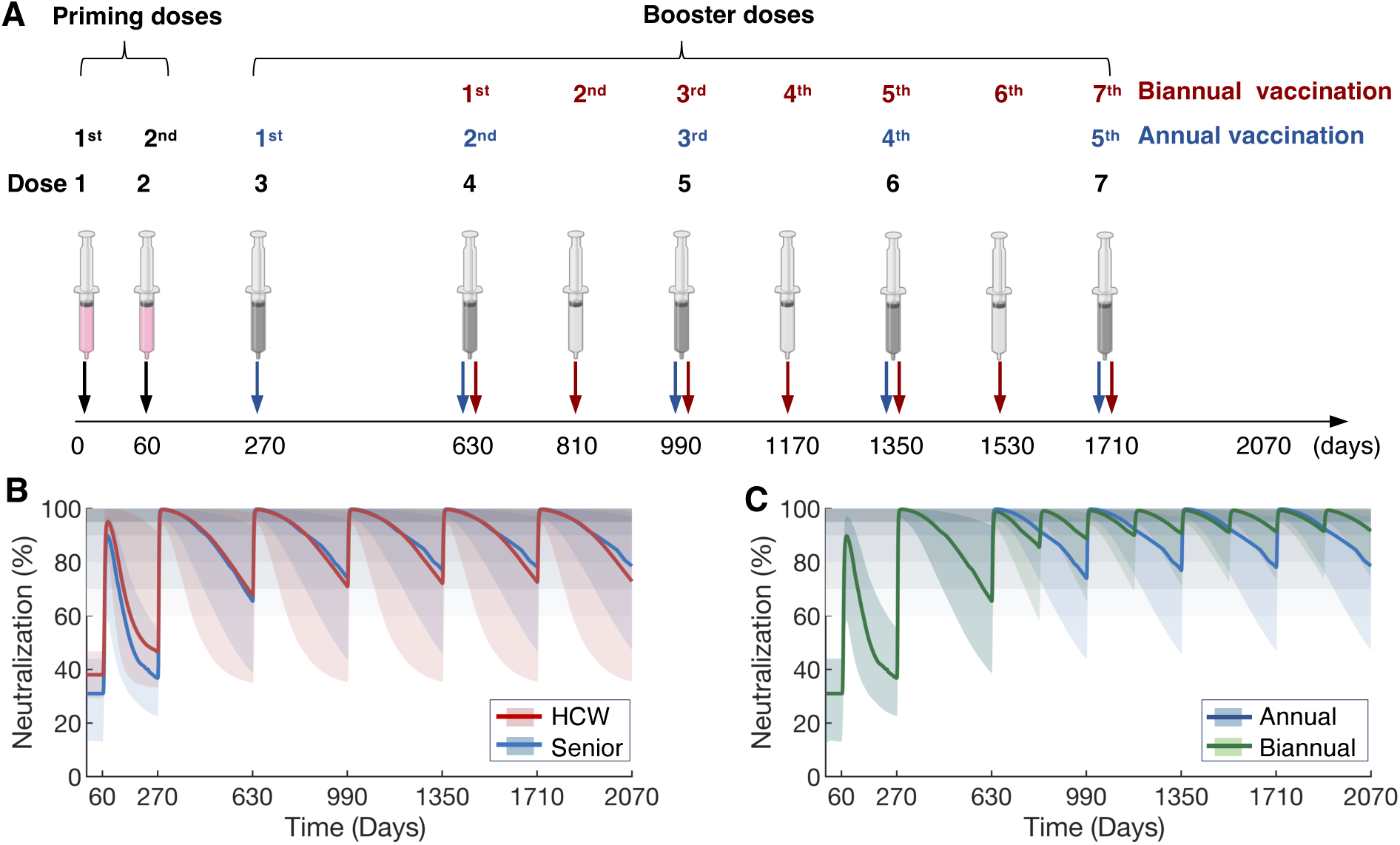
Predicted neutralization dynamics for annual and biannual booster campaigns. **A)** Schedule of annual and biannual boosters. Pink doses represent the two full priming doses, while dark doses indicate annual booster doses (full or half doses), and light gray doses represent additional biannual doses. Black arrow: administration of primary doses. Blue arrow: administration of annual boosters. Red arrow: biannual boosters starting one year after the first booster. **B)** Predicted neutralization for HCWs and seniors. Here, healthcare workers received an annual half-dose booster, while seniors received an annual full-dose booster. Red solid line: median neutralization in the HCW cohort. Blue solid line: median neutralization in the senior cohorts. Shaded regions: 95% confidence intervals for the estimated neutralization outcomes in respective cohorts. **C)** Predicted neutralization curves for seniors receiving annual full boosters versus biannual half-dose boosters. Blue solid line: median neutralization after annual full booster. Green solid line: median neutralization after biannual half-dose booster. Shaded regions: 95% confidence interval of the predicted neutralization for each cohort. Grey horizontal bands: neutralization thresholds of 60%, 70%, 80%, 90%, and 95%.

If the entire cohort were to receive annual full boosters, our model predicted that neutralization levels would steadily increase over time, largely tapering off after the third booster (fifth vaccine dose). The HCW cohort was predicted to exhibit superior vaccine responses, maintaining slightly higher median neutralization levels **(Supplementary Figure 7A)**. Specifically, half of HCWs maintained neutralization levels above 70% throughout the year following the second booster, with median levels exceeding 80% for HCWs and reaching 90% by the fourth booster, while for seniors, median levels remained between 70% and 80%. **(Supplementary Figure 7C**). If younger individuals were to continue receiving annual half-dose boosters with seniors administered full-dose boosters, neutralization levels for HCWs slightly increased, with a median of about 70%. Seniors, however, were predicted to achieve higher neutralization levels compared to HCWs, with median levels around 75% after the second booster, approaching 80% thereafter (**Figure 5B** and **Supplementary Figure 7D**).

As expected, providing annual half-dose boosters to seniors was not predicted to generate strong protection. Half-dose boosters for seniors resulted in a continuous decline in neutralization levels, with median trough levels falling below 60% before the administration of subsequent boosters (**Supplementary Figures 7B and E**). Thus, we used our model to investigate whether more frequent half-dose boosters could maintain high neutralization between vaccine doses. We found this strategy to significantly increase neutralization outcomes in seniors, with noticeable improvements prior to receiving the next booster 6 months later (**Figure 5C**). Compared to annual full-dose vaccination, our model predicts that biannual half-dose vaccination provides less durable neutralization during the first six months but significantly improves in the second half of the year with the administration of the half-booster. Remarkably, after just two biannual half-dose boosters, seniors were predicted to maintain a median neutralization level of about 90% and keeps slightly increasing before the next vaccination (**Figure 5C and Supplementary Figure 7F**).

## DISCUSSION

mRNA COVID-19 vaccines have proven highly effective in preventing severe illness and reducing mortality by inducing robust cellular immune responses. However, contraction of humoral immunity, particularly antibody responses, increases the risk of reinfection and severe illness, especially in seniors who have generally weaker vaccine responses[13]. To design vaccination schedules that account for waning immunity and ensure long-term humoral population-level protection, it is important to understand this age-related disparity. To this end, we developed a mechanistic mathematical model that describes the initiation and establishment of the humoral response following vaccination, and the recall of B cell memory during repeated vaccination. Through calibration to clinical data from healthcare workers and seniors who received the three-dose primary mRNA COVID-19 vaccine series (two priming doses and a booster)[13], our model successfully captured the variability in antibody responses by T follicular helper cell decay and vaccine-induced immunological amplification parameters. Specifically, our results revealed a strong correlation between age and Tfh cell decay rates (**Figure 2C–D**), with seniors a median rate of decay nearly twice as fast as that of HCWs. Due to the rapid elimination of Tfh cells, seniors were less able to activate B cells, the primary source of antibody production, leading to lower antibody concentrations. Repeated vaccination was found to continuously enhance (with diminishing returns) immunological amplification with respect to the activation of naïve B cells by Tfh cells and the acceleration of antibody production rates by both plasmablasts and long-lived plasma cells. Notably, we found no statistically significant age-related differences in these amplification factors, suggesting that differences in cellular populations and functional responses drive age-related differences in vaccination outcomes.

We used model-predicted neutralization outcomes to compare vaccine-induced immunity between age groups, given that viral neutralization is key to infection preventions. Our analyses showed no statistically significant difference between the two age groups with respect to key neutralization parameters (i.e., half baseline-maximal effect or sensitivity of neutralization levels to antibody concentrations). However, seniors were predicted to have lower neutralization baseline due to weaker response to the first dose compared to HCWs (**Supplementary Figure 3C–E**). To evaluate vaccine performance, we analyzed neutralization levels and the likelihood of maintaining neutralization above thresholds of 60%, 70%, 80%, 90%, and 95%. Our results show that healthcare workers were more likely maintain neutralization across these four thresholds while also maintaining neutralization above them for longer after the two full priming doses. However, since the HCWs in the clinical study used to calibrate our mathematical model only received a half-dose booster, the neutralization profiles of seniors (who received a full third dose) become comparable to those of healthcare workers after the booster dose. We confirmed this dose dependency by simulating full-dose boosters for HCWs, which showed that HCWs would achieve higher antibody, long-lived plasma cell, and memory B cell concentrations compared to seniors using this strategy.

We then hypothesized that dose sizes could be adjusted to improve neutralization dynamics. The predicted neutralization patterns after mixed-dose sequences highlight the importance of receiving a full dose for the final administration in a three-dose schedule, contrary to the regimen administered to healthcare workers in our cohort. In fact, our model predicted that healthcare worker could even achieve higher neutralization with two half primary doses and a full booster, further underlining the importance of booster dose size. Based on these results, we explored how to best achieve higher neutralization in annual vaccination campaigns. We found that annual half-dose boosters could provide HCWs with slightly improved neutralization profiles. However, seniors would need annual full-dose boosters to achieve neutralization profiles comparable to those after the third dose. To further enhance neutralization responses in seniors, biannual half-dose boosters could be used to provide higher protection against infection.

There are several limitations to our study. We did not account for the impact of vaccine/variant mismatch, given that the data we used for model calibration was collected during different waves of COVID-19 (e.g., Delta, Omicron). With additional data, we could explore vaccine efficacy against different variants and use our model to study vaccination strategies with respect to neutralization thresholds for different variants. Moreover, our neutralization function (Eq. 14) required adjustment from the usual dose-response relationship due to the limited accuracy of having only a single neutralization measurement during the interval between the initial two doses. Thus, we introduced a baseline neutralization to quantify vaccine-induced neutralization after the primary dose. Under this assumption, neutralization levels cannot fall below this baseline even over extended periods, which limits the time over which our predictions remain valid. Lastly, in our data, all antibody concentrations exceeding the upper limit of quantification (ULQ) were recorded as the ULQ, which could lead to estimation errors, particularly when assessing the amplification factor after the third dose. However, when estimating parameter values, we applied censoring data methods, allowing the estimated antibody curves to extend beyond the ULQ. While this approach could impact on the accuracy of our parameter estimates, it remains a valid and appropriate method for handling such data.

This study elucidates age-related differences in antibody kinetics, particularly the accelerated decay of T follicular helper cells, and establishes benchmarks for ongoing mRNA COVID-19 vaccination in both younger and older adults. Thus, this work highlights the value of mechanistic mathematical modelling for understanding heterogeneity in vaccine-induced responses and tailoring vaccination strategies for different demographic groups. Ultimately, our mathematical framework is critical to optimize vaccination schedules while enabling the development of cost-effective, personalized medical interventions that are critical for combating infectious diseases, which can be challenging to assess in real time.

## CRediT authorship contribution statement

**Xiaoyan Deng**: Conceptualization, Data curation, Formal analysis, Investigation, Methodology, Writing–original draft, Writing–review&editing. **Suzan Farhang-Sardroodi**: Investigation, Methodology, Writing–review & editing. **Morgan Craig**: Conceptualization, Resources, Funding acquisition, Investigation, Methodology, Supervision, Writing–review & editing.

## Acknowledgements

The authors would like to thank the COVID-19 Immunity Taskforce (CITF) for facilitating the data sharing that made this study possible. We also sincerely thank all the patients, family members, and staff who participated in the original study.

## Data and code availability

Data requests should be directed to the COVID-19 Immunity Taskforce Databank, available at https://www.covid19immunitytaskforce.ca/citf-databank/. All code for reproducing figures in the this study is available on Github: https://github.com/Craig-Lab/COVID19Vaccine.git.

