## Supplementary for "Predicting age-related determinants of heterogeneous outcomes to COVID-19 mRNA vaccines through mathematical modelling"

***Parameter estimation***

Parameter values were estimated from various sources, including previous studies, and stepwise fitting of the timing to peak values of specific outputs, detectable period, or the relative magnitude of biomarkers and average antibody data of the entire cohort.

**1. Estimation according to mean lifetime or half-life:**

To calculate the decay rate of lipid nanoparticles, cells, and antibodies, we used the standard half-life relationship (Eq. S1):

$$t_{\frac{1}{2}} = \tau ln2 = \frac{ln2}{\lambda}, \qquad \text{(S1)}$$

where $t_{\frac{1}{2}}$ is the half-life, $\tau$ represents mean life, and $\lambda$ is the rate of decay.

1.1 LNP degradation rate ($d_L$)

Lipid nanoparticles have an estimated the mean life up to 10 days[1–3]. According to the relationship between mean life and decay constant (Eq.S1):

$$d_L = \frac{1}{10\text{ days}} = \frac{0.1}{day}.$$

1.2 Absorption rate LNPs by vaccinated cells ($\delta_{LV}$)

Pardi, N. *et al.* [10] estimated the active translation of mRNA in the liver for 1-4 days. Based on this, we assumed the mean duration of mRNA translation to be 4 days and considered the total decay of lipid nanoparticles ($d_L + \delta_{LV}$) to be the decay of mRNA molecules:

$$d_L + \delta_{LV} = \frac{1}{4\text{ days}} = \frac{0.25}{day}.$$

We then calculated $\delta_{LV}$ by substracting the estimated $d_L$ value from section 1.1:

$$\delta_{LV} = (0.25 - 0.1) = \frac{0.15}{day}.$$

1.3 Natural death rate of activated B cells ($d_B$)

Kogut *et al.*[4] stated that the half-life of activated B cells ranges from 5-6 weeks. We chose 5 weeks (35 days) as the half-life, which implies that

$$d_B = \frac{\ln(2)}{35\text{ days}} \approx \frac{0.02}{day}.$$

1.4 Degradation rate of IL-21 ($d_I$)

Schmidt, H. *et al.*[5] estimated the mean half-life of IL-21 ranges from 3.6 to 5.3 hours We selected a half-life of 5 hours, which corresponds to 0.2 days. Using this, the decay rate ($d_I$) was calculated per Eq. S1:

$$d_I = \frac{\ln(2)}{0.2\text{ days}} \approx \frac{3.47}{day}.$$

1.5 Natural death rate of long-lived plasma B cells *($d_P$)*

Koike, T. *et al.* [6] estimated the half-life of IgG plasma cells in the bone marrow to be 61.8 days, so we calculated the decay rate for plasma cells as:

$$d_P = \frac{\ln(2)}{61.8\text{ days}} \approx \frac{0.0112}{\text{day}}.$$

1.4 Natural death rate of memory B cells ($d_M$)

Dan, J. M. *et al.*[7] reported no apparent half-life at 5 to 8 months post-infection, while other studies estimated the half-life to be 8-10 weeks[8]. We chose the half-life to be 8 months (240 days), and calculated the decay rate for memory B cells to be:

$$d_M = \frac{\ln(2)}{240\text{ days}} \approx \frac{0.0029}{day}.$$

1.5 Antibody degradation rate ($d_A$)

Dan, J. M. *et al.*[7] estimated the half-life of IgG in the bone marrow to be 21 days, and we used it to approximate the antibody decay in our model, giving:

$$d_A = \frac{\ln(2)}{21\text{ days}} \approx \frac{0.033}{day}.$$

2. **Estimation according to cell cycle**:

Cell doubling time represents the time required for a population or quantity to double in size under exponential growth and is given by

$$t_{double} = \frac{ln2}{\lambda}, \qquad \text{(S2)}$$

where $\lambda$ is the exponential growth rate.

2.1 Proliferation rate of GC B cells ($\beta_G$)

The cell cycle of GC B cells has been estimated to range between 4 and 6 hours[9,10]. Here, we chose 6 hours (equivalent to 0.25 days) as the GC B cell cycle. Therefore, we calculated the proliferation rate of GC B cells using Eq. S2 to obtain:

$$\rho_G = \frac{\ln(2)}{0.25\text{ days}} \approx \frac{2.77}{day}.$$

3. **Estimation based on time to peak:**

To estimate the rates based on the peak time of cells, we performed nonlinear least-square curve fitting using the *lsqnonlin* function in Matlab R2022a. The objective function (Eq. S3) minimized the difference between the estimated peak time from the model ($\hat{t}_{peak}$) and the target peak time ($t_{peak}$) chosen from literature:

$$objective = \left(\hat{t}_{peak} - t_{peak}\right)^2 \qquad \text{(S3)}$$

3.1 Decay rate of T follicular helper cells ($d_T$) and Tfh activation rate by vaccinated cells ($\delta_{TV}$)

T follicular helper (Tfh) cells have been shown to peak between days 4 and 10[11–14] after vaccination. Based on this, and using a target peak time of 4 days. we estimated two parameters in Eq. 3 of the main text: the Tfh activation rate by vaccinated cells ($\delta_{TV}$) and the decay rate of Tfh cells ($d_T$).

By solving the optimization problem (Eq. S3), we obtained

$$\delta_{TV} = 0.73 \text{ cells/AU/mL and } d_T = \frac{0.57}{day}.$$

3.2 Decay rate of vaccinated cells ($d_V$)

Given a target peak time of 4 days for Tfh cells, we assumed that vaccinated cells peak 2 days earlier to the peak of Tfh cells (i.e., day 2). Vaccine mRNA can be detected in blood up to 28 days[15,16], hence we set the detectable period to be 28 days. Therefore, the objective function in this case can be expressed as:

$$objective = \left(\hat{t}_{peak} - t_{peak}\right)^2 + (V(28) - \epsilon)^2,$$

where $V(28)$ is the concentration of vaccinated cells at day 28, and $\epsilon = 10^{-6}$ is a small threshold value representing undetectable levels of vaccinated cells at day 28. We thus obtained the decay rate of vaccinated cells of

$$d_V = \frac{0.89}{day}.$$

3.3 B-cell activation rate by Tfh cells ($\delta_{BT}$ and $(\rho_S + \rho_G)$)

Studies[17–19] report that the peak time for short-lived plasmablasts ranges between 6 and 12 days. Considering the sequential process of Tfh cell stimulation, B cell activation by Tfh, and plasmablast generation, we assumed the peak time for B cells to be 5 days and the peak time for plasmablasts to be 8 days. By solving Eq. S3, we estimated $\delta_{BT}$ and the sum of the generation of plasmablasts and GC B cells ($\rho_S + \rho_G$) to be

$$\delta_{BT} = \frac{0.36}{day},$$

and

$$(\rho_S + \rho_G) = \frac{1.29}{day}.$$

3.4 Generation rate of plasma blasts by activated B cells ($\rho_S$) and death rate of plasmablasts ($d_s$)

We used 8 days as the peak time for plasmablasts, midway between the estimated 6-12 days range[17–19]. We fit the two parameters in plasmablasts $\rho_S$ and $d_s$ (Eq. 7 in Main Text) and obtained:

$$\rho_S = \frac{0.60}{day} \text{ and } d_S = \frac{0.28}{day}.$$

Since we have the estimation for the sum $(\rho_S + \rho_G) = \frac{1.29}{day}$ from section 3.3, therefore,

$$\rho_G = \frac{0.69}{day}.$$

3.5 GC B cells stimulation rate by IL-21 ($\delta_{IG}$) and natural death rate of GC B cells ($d_G$)

The estimated peak time for GC B cells ranges from 6 to 12 days[11,20,21]. We chose 7 days for the peak time and estimated the two rates stimulation of germinal center B cells by IL-21and the natural death rate of GC B cells. This gave

$$\delta_{IG} = 0.73 \text{ and } d_G = \frac{0.59}{day}.$$

3.6 Differentiation probability of GC B cells into LLPCs ($p_{p1}$)

Hoyer *et al.* estimated the proportion of short-lived and long-lived plasma cells in mice with chronic humoral autoimmunity. They found that approximately 60% of antibody-secreting cells are short-lived plasma cells, while 40% are long-lived plasma cells within 10 days. To capture this relative distribution, we fit the differentiation probability of GC B cells into LLPCs ($p_{p1}$) such that the area under the curve (AUC) for plasmablasts up to day 10 is 1.5 times the AUC of the long-lived plasma cells, i.e., $1.5 \times AUC_{LLPC}(10) = AUC_{PB}(10)$, giving

$$p_{p1} = 0.24.$$

3.7 Production rate of antibody by long-lived plasma cells ($\alpha_P$) and plasmablasts ($\alpha_S$)

The peak time of antibody production ranges between 18 and 40 days[22–24]. The average antibody concentration from Mwimanzi *et al.*[25] measured on day 30 after the first vaccination, was 88.86 AU/ml. We chose 21 days as the peak time and estimated production rates by minimizing the sum of squares:

$$objective = \left(\hat{t}_{peak} - t_{peak}\right)^2 + (A(21) - 88.86)^2,$$

where $A(21)$ is the model predicted antibody concentration. This gave

$$\alpha_P = 271 \text{ AU/cells/day and } \alpha_S = 18 \text{ AU/cells/day}.$$

4. **Estimation for the remaining parameters**

4.1 Half maximal effect of IL-21 ($EC_{50,I}$)

We estimated this values as half of the maximum IL-21 concentrations after the first dose, which yields:

$$EC_{50,I} = \frac{\max(I)}{2} = 0.03\, pg/mL.$$

4.2 Probability of memory B cell self-renewal ($p_M$)

To estimate this probability, we considered the immune response following the second vaccine dose. To determine the self-renewal probability of memory B cells, we used estimates reporting that germinal center persists at least 14 days after immunization[26–28]. We assumed their durability extends up to 30 days after the second dose. Similar to the fitting process for vaccinated cells, we minimized the function

$$(B_G(30) - \epsilon)^2,$$

where $B_G(30)$ represents the concentration of germinal center B cells at day 30, and $\epsilon = 10^{-6}$ is a small threshold value representing undetectable levels of germinal center B cells at day 30. Therefore, we have $p_M$= 0.85.

***Neutralization can be further boosted by delaying the second in seniors***

We studied the impact of dose size on vaccination outcomes and sought to determine whether neutralization in seniors could be further improved by optimizing the three-dose primary vaccination series. For this, we adjusted the dosing intervals reported by Mwimanzi *et. al*[25] and assumed seniors received all three vaccines according to the guidelines in place at the time of the study. Specifically, we varied the interval between the first and second dose from 1, 2, 4 and 6 months, keeping a 7-month gap before the booster (**Supplementary Figure 8A**), or maintained a 2-month interval between the initial two doses and varied the timing of the third dose from 6 to 9 months (**Supplementary Figure 8B**). The analysis covered the same timeframe as previously described, from initial vaccine administration to one year after the third dose (0 to 630 days) based on the current vaccination regimen. Note that day 630 may not correspond exactly to one year after completing the three-dose series due to these adjustments in dose intervals.

Overall, our model predicted that the time between doses in the primary vaccination series did not significantly affect peak neutralization levels but shifted the curves rightward over time, thereby enhancing final neutralization level by day 630 (**Supplementary Figure 8C–D**). Notably, delaying the second dose proved slightly more effective than postponing the booster in enhancing final neutralization levels. Comparing two regimens that extend either the first or second dosing interval by 2 months while keeping the other interval fixed, our results indicate that a 4-month gap between the first two doses leads to slightly higher neutralization levels in seniors compared to a 9-month gap between the second and third doses, with median levels of 75.28% and 74.45%, respectively (**Supplementary Figure 8C–D**). Indeed, delaying the second dose to six months after initial vaccination in seniors increased the median neutralization from 67.43% to 82.57% (**Supplementary Figure 8E)**. This delay significantly improved neutralization outcomes at the 90% threshold, extending the median duration by 61 days (from 515 to 576 days) and increasing the number of seniors maintaining 90% neutralization on day 630 fivefold, from 2 to 10 (**Supplementary Figure 8F**). Additionally, four seniors achieved over 95% neutralization on day 630, whereas no seniors could maintain this level under the current regimen (**Supplementary Figure 8G**).

**Supplementary Table 1. Model parameter values.**

| Variable | Parameter | Definition | Value | Units | Reference |
|---|---|---|---|---|---|
| Lipid nanoparticle (LNPs; L) | $d_L$ | LNP degradation rate | 0.1 | 1/day | fitted |
| Vaccinated cells (V) | $d_V$ | Death rate of vaccinated cells | 0.89 | 1/day | fitted |
| | $\delta_{VL}$ | Generation rate of vaccinated cells by LNPs | 0.15 | 1/day | fitted |
| T follicular helper cells (Tfh; $T_h$) | $\delta_{TV}$ | Tfh cell activation rate by vaccinated cells | 0.73 | cells/AU/mL | fitted |
| | $d_T$ | Decay rate of T follicular helper cells | 0.57 | 1/day | fitted |
| Activated B cells ($B$) | $\delta_{BT}$ | B-cell activation rate by Tfh | 0.36 | 1/day | fitted |
| | $d_B$ | Natural death rate of activated B cells | 0.02 | 1/day | [20,29] |
| Germinal center B cells (GC B; $B_G$) | $\rho_G$ | Generation rate of GC B cells by activated B cells | 0.69 | 1/day | fitted |
| | $\beta_G$ | Proliferation of GC B cells | 2.77 | 1/day | fitted |
| | $\delta_{IG}$ | Stimulation of germinal center B cells by IL-21 | 0.73 | – | fitted |
| | $p_G$ | Probability of self-renew of GC B cells | 0.1 | – | [30] |
| | $d_G$ | Natural death rate | 0.59 | 1/day | fitted |
| Short-lived plasmablasts (SLPBs; $P_S$) | $\rho_S$ | Generation rate of SLPBs by activated B cells | 0.60 | 1/day | fitted |
| | $d_S$ | Death rate of SLPBs | 0.28 | 1/day | fitted |
| Long-lived plasma cells ($LLPCs$; $P_L$) | $p_{p1}$ | Differentiation probability of GC-B cells into LLPCs | 0.26 | – | fitted |
| | $p_{p2}$ | Differentiation probability of memory B cells to LLPCs | 0.9 | – | [31] |
| | $d_P$ | Natural death rate of plasma B cells | 0.0112 | 1/day | [6] |
| Memory B cells ($M$) | $\beta_M$ | Proliferation of memory B cells | 0.009 | 1/day | [32] |
| | $p_M$ | Probability of self-renew of memory B cells | 0.85 | – | fitted |

| | | | | | |
|---|---|---|---|---|---|
| | $d_M$ | Natural death rate | 0.0029 | 1/day | [7] |
| Antibodies ($A$) | $\alpha_P$ | Production rate of antibody by long-lived plasma cells | 18 | AU/cells/day | fitted |
| | $\alpha_S$ | Production rate of antibody by short-lived plasma cells | 271 | AU/cells/day | fitted |
| | $d_A$ | Antibody degradation rate | 0.033 | 1/day | [7] |
| Interleukin 21 (IL-21; $I$) | $\rho_I$ | Production rate of IL-21 by Tfh cells | 2.07 | 1/day | [33] |
| | $EC_{50,I}$ | Half maximal effect of IL-21 | 0.03 | pg/mL | fitted |
| | $d_I$ | Degradation rate of IL-21 | 3.47 | 1/day | fitted |

**Supplementary Table 2. AIC and BICs of the model simulated in the cohort.**

| Fitted parameters | AIC | BIC |
|---|---|---|
| $d_t$ and $\delta_{BT}$ | 5628.59 | 5645.21 |
| $d_t$ and $\alpha_S$ | 6208.22 | 6224.84 |
| $d_t$, $\delta_{BT}$, and $\alpha_S$ | 6403.06 | 6419.68 |
| $d_t$, $\delta_{BT}$, $\alpha_S$, and $\alpha_P$ | 5656.44 | 5673.06 |

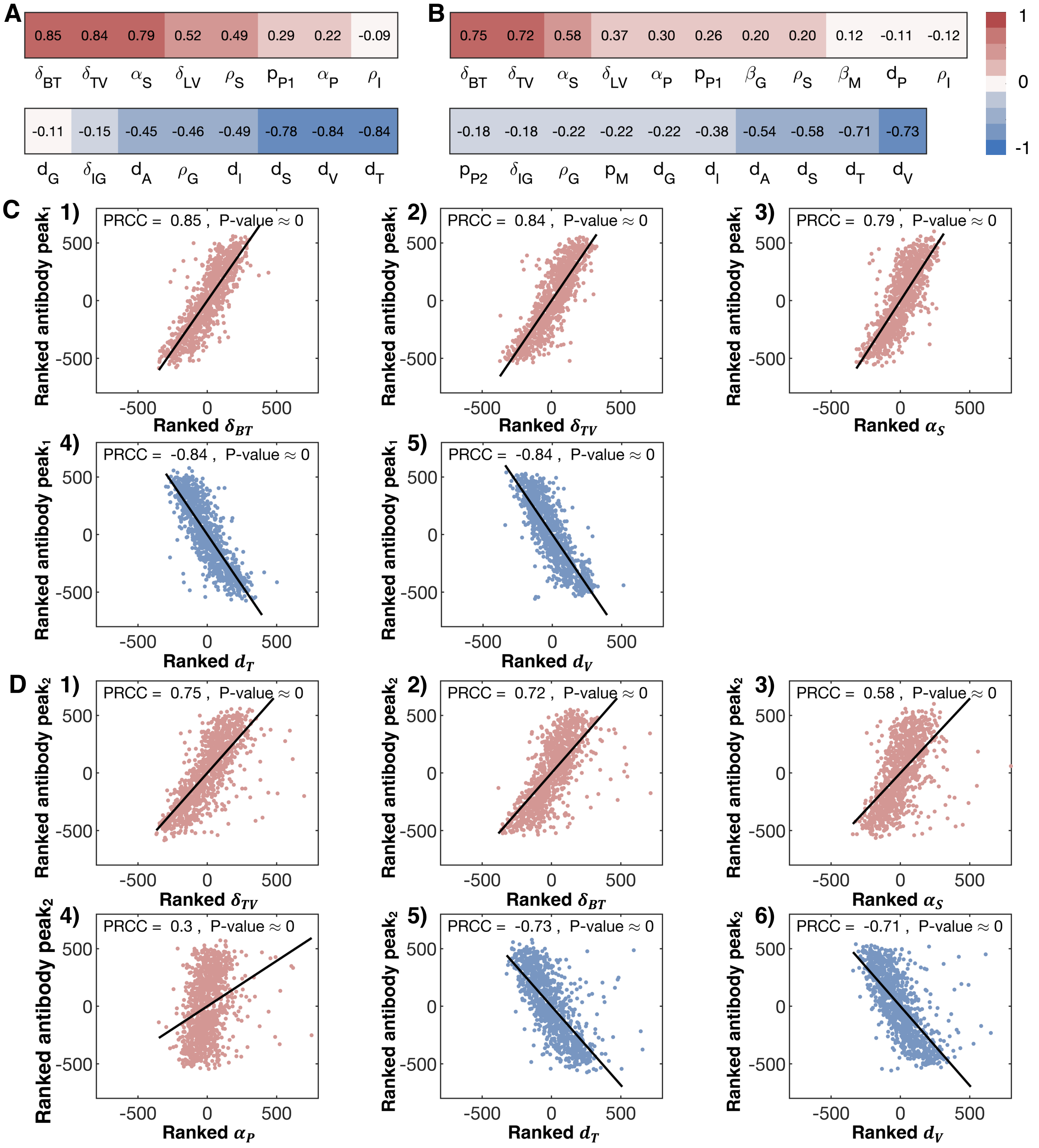


**Supplementary Figure 1. Partial rank correlation coefficient plots. A-B)** PRCC plot showing all statistically significant parameters (p-value < 0.05) with respect to peak antibody concentrations after the **A)** first and **B)** second primer dose. Red squares: positive PRCC. Blue squares: negative PRCCs **C–D)** PRCC scatter plot of ranked input parameter and ranked antibody peaks after the **C)** first and **D)** second primer dose. $d_T$: Th cell decay rate; $d_V$: death rate of vaccinated cells; $\delta_{BT}$: generation rate of activated B cells by Th cells; $\delta_{TV}$: production rate of Th cell stimulated by vaccinated cells; $\alpha_S$ : antibody production rate by short-lived plasmablasts; $\alpha_P$ : antibody production rate by long-lived plasma cells (panel **D**). Solid black line: fitted regression between ranked antibody peak values and corresponding parameter. Blue filled circles: seniors. Red filled circles: HCWs.

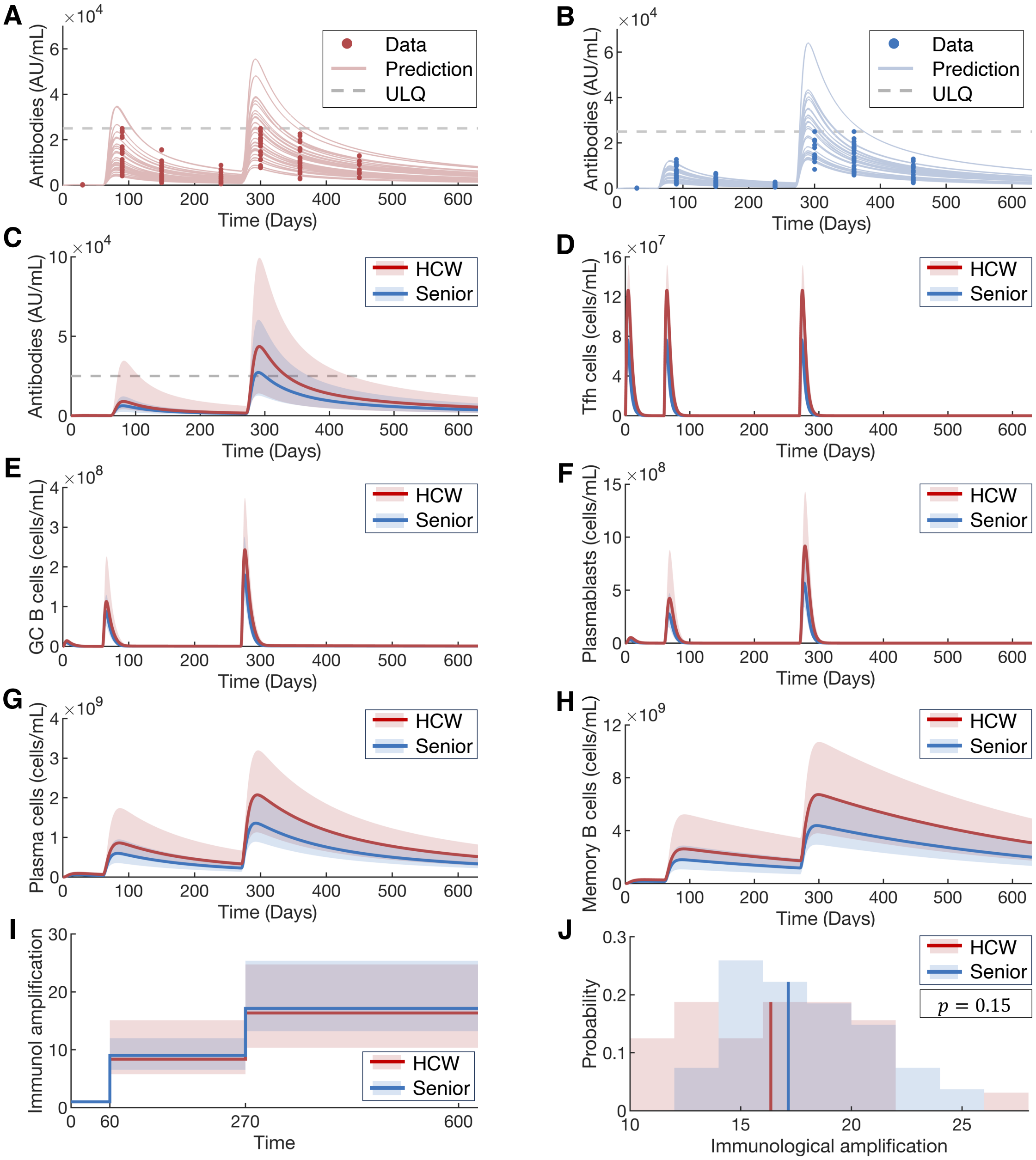


**Supplementary Figure 2. Predicted dynamics of antibody and immunological biomarkers. A)** Model predicted antibody concentrations in healthcare workers. **B)** Model predicted antibody dynamics in seniors. Dots: measured antibody concentrations. **A–B)** Solid lines: individual model predicted antibody dynamics. Gray dashed lines: upper limit of quantification (ULQ). **C–H)** Predicted antibody, T follicular helper cells, germinal center B cells, plasmablasts, long-lived plasma cells, and memory B cells dynamics for both seniors (blue) and HCWs (red). In each, HCWs were simulated to receive a full booster (third) dose. **I)** Immunological amplification over time, with vaccination on days 0, 60, and 270. **J)** Distribution of cumulative immunological amplification after all three doses. Vertical lines: median values. Indicated p-values are from a Wilcoxon-Mann-Whitney test with $\alpha = 0.05$ level of significance. **C–I)** Solid lines: predicted median antibody and neutralization levels. Shaded areas: 95% confidence interval based on individual predicted curves. **A–J)** Blue filled circles: seniors. Red filled circles: HCWs.

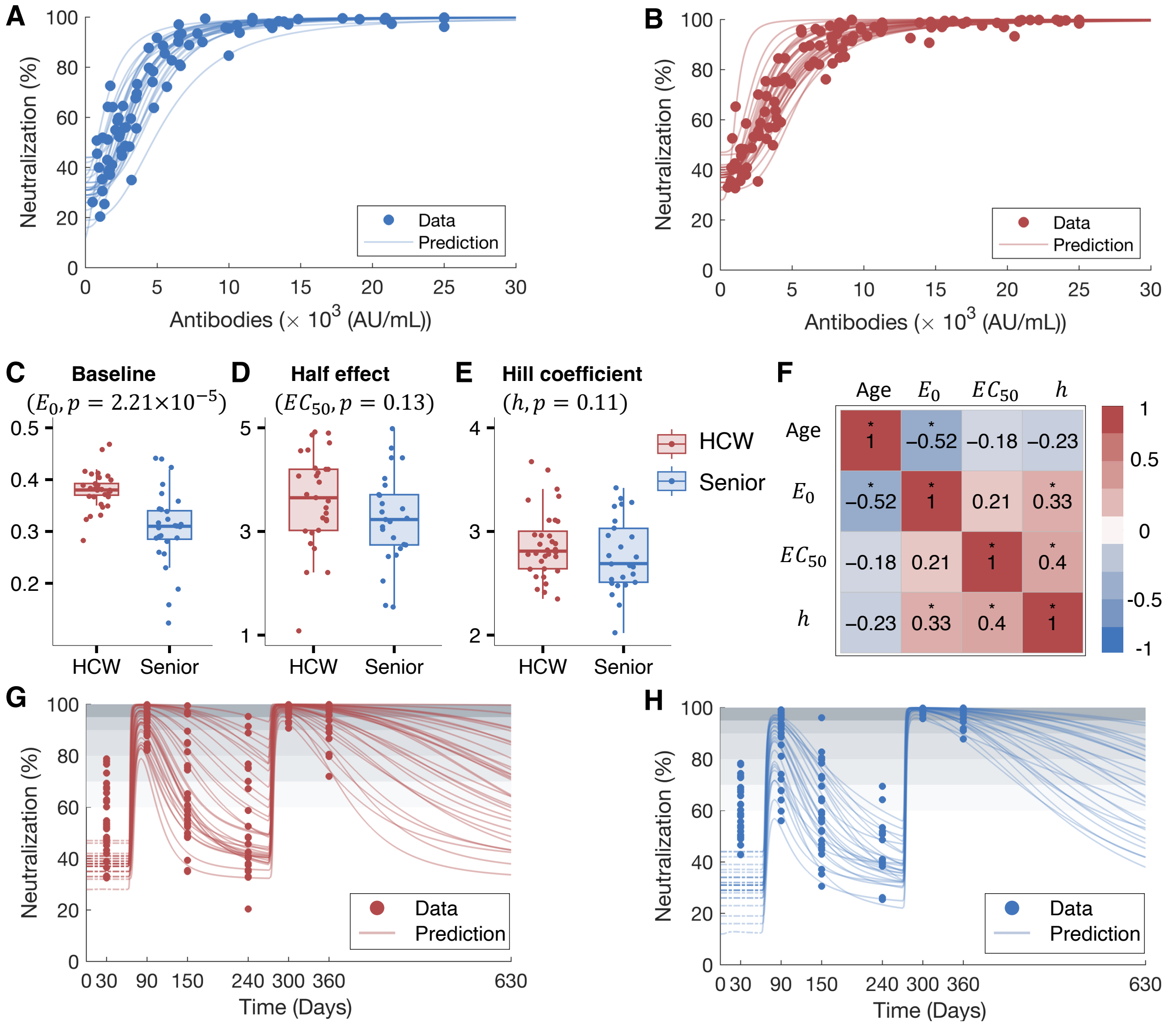


**Supplementary Figure 3. Estimated parameters for the antibody-neutralization association and predicted neutralization curves. A)** Fitted antibody-neutralization curves for HCWs. **B)** Fitted antibody-neutralization curves for seniors. Boxplots of **C)** baseline neutralization ($E_0$), **D) D)** half-maximal effective concentration ($EC_{50}$), and **E)** Hill coefficient ($h$) for the antibody-neutralization relationship (Eq. 14 in the Main Text). **F)** Correlation matrix between estimated parameter values and age. Pearson correlation values are indicated by the size and value of the squares, with '*' denoting statistical significance at the 5% levels. **G)** Predicted individual neutralization patterns for HCWs. **H)** Predicted individual neutralization patterns for seniors. **A–B)** and **G–H**) Blue filled circles: senior data[25]. Red filled circles: healthcare worker data[25]. Solid lines: model-predicted antibody and neutralization dynamics for each individual. **G–H)** Shaded regions: neutralization levels of 60-70%, 70-80%, 80-90%, 90-95%, and above 95% from light to dark gray, respectively.

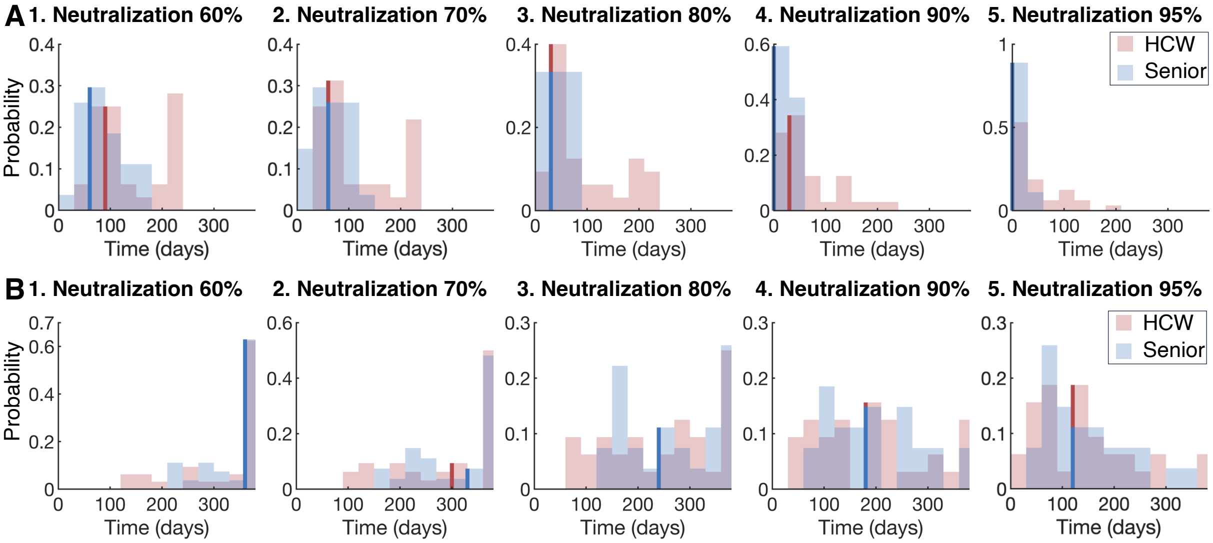


**Supplementary Figure 4. Distribution of predicted duration above neutralization threshold after the second and third vaccinations. A)** Distribution of predicted duration above neutralization thresholds 60% to 95% after the second dose. **B)** Distribution of predicted duration above neutralization thresholds 60% to 95% after the booster. Blue bars: seniors. Red bars: healthcare workers. Vertical lines: median value (i.e., 50% of the population)

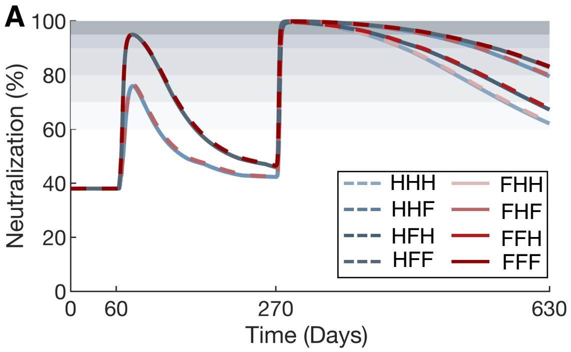


**Supplementary Figure 5. Predicted average antibody response and neutralization dynamics with varying dose sizes. A)** Predicted neutralization response for HCWs with different combination of half-and-full doses. **B)** Predicted neutralization response for seniors with different combination of half-and-full doses. **A–B**) Blue dashed line: three-dose regimens starting with a half dose. Red solid lines: regimens starting with a full dose. "H": half dose. "F": full dose. From light to dark gray, the shaded regions represent neutralization levels of 60-70%, 70-80%, 80-90%, 90-95%, and above 95%, respectively.

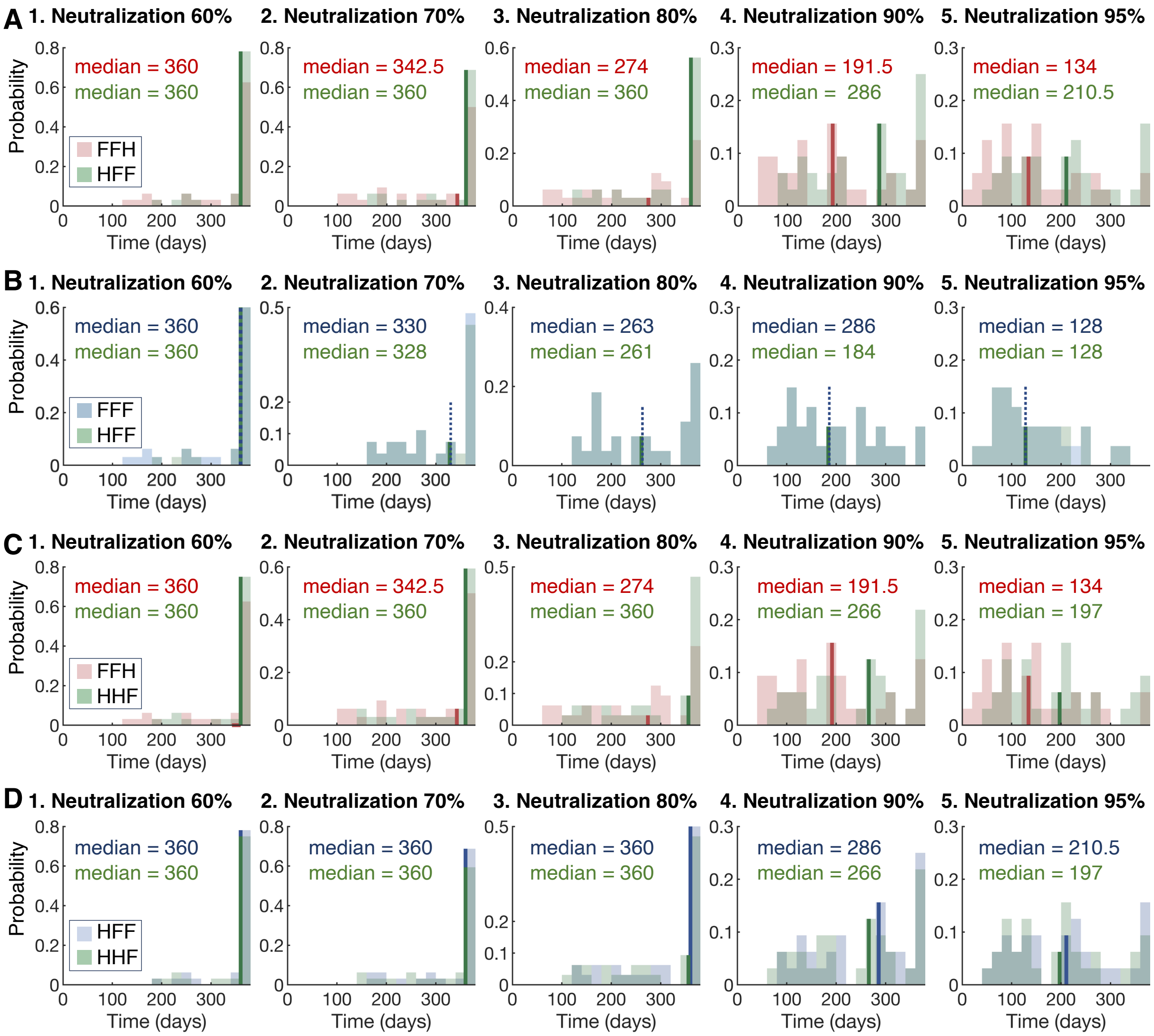


**Supplementary Figure 6. Predicted duration above neutralization threshold (60% to 95%).** **A)** Comparison of predicted "FFH" (current vaccination strategy for HCWs) and "HFF" (half priming doses followed by two full doses) strategies for healthcare workers. **B)** Comparison of predicted “FFF” (current vaccination strategy for seniors) and “HFF” strategies in seniors. **C)** Comparison of “FFH” and “HHF” strategies (two half priming doses followed by a full booster) in HCWs. **A–C**) Red bars: seniors under current vaccination strategy. Blue bars: healthcare workers under current vaccination strategy. Green bars: each cohort under improved vaccination strategy. **D)** Comparison of “HFF” and “HHF” strategies in HCWs. Blue bars: predicted distributions for HCWs under the “HFF” and “HHF” strategies, respectively. **A–D)** Vertical lines median values. “H”: half dose. “F”: full dose.

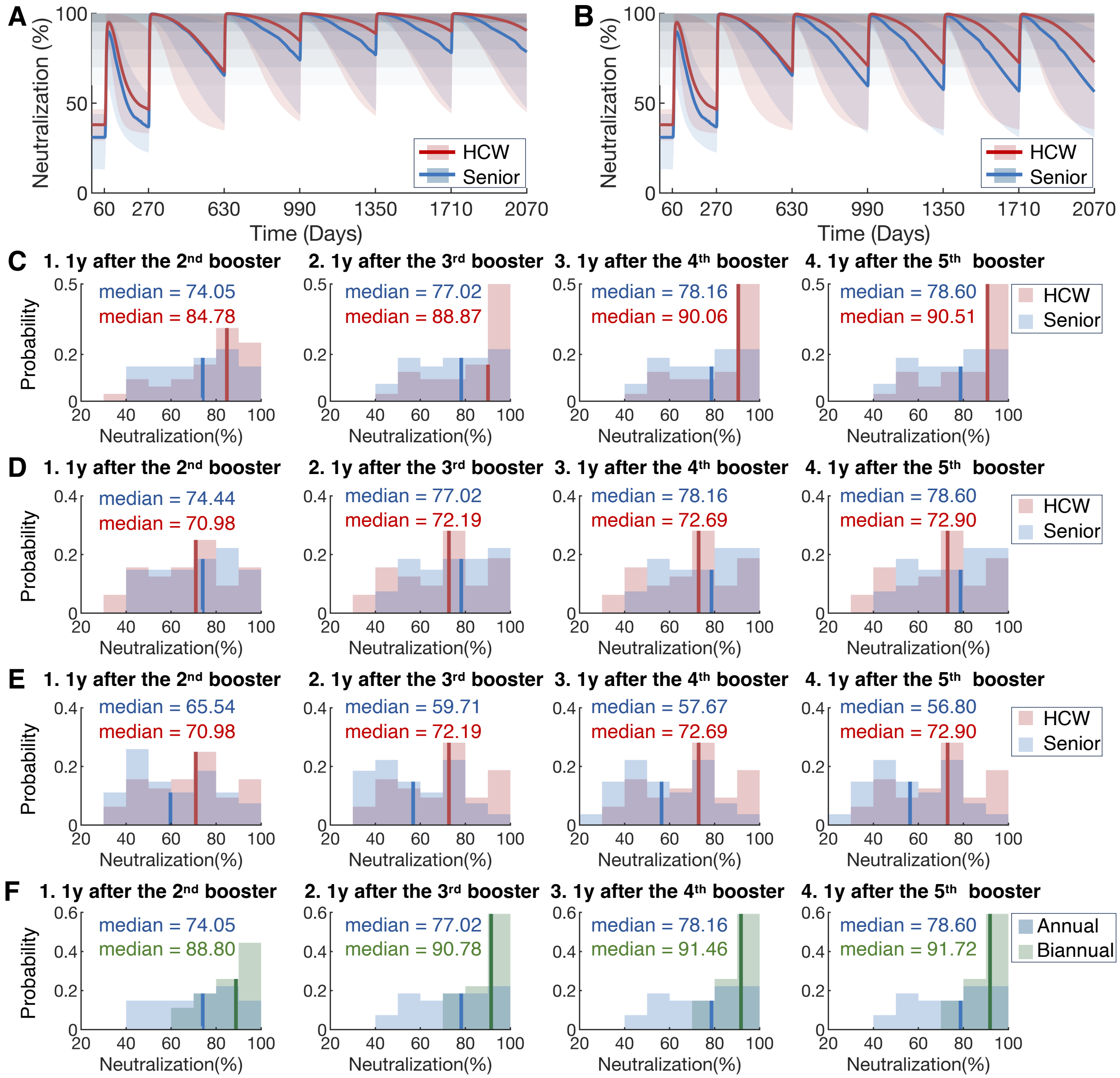


**Supplementary Figure 7. Predicted neutralization patterns after annual and biannual boosters and neutralization levels prior to next booster. A)** Predicted neutralization curves for seniors and healthcare workers after receiving an annual full-dose booster. **B)** Predicted neutralization curves for seniors and healthcare workers receiving an annual half-dose booster. Blue solid lines: median predicted neutralization for seniors. Red solid lines: median predicted neutralization HCWs. Shaded regions: 95% confidence interval based on estimation of individual predictions. From light to dark gray: neutralization levels of 60-70%, 70-80%, 80-90%, 90-95%, and above 95%, respectively. **C)** Distribution of neutralization levels one year after full dose boosters 2 to 5. **D)** Distribution of neutralization levels one year after boosters 2 to 5 under current vaccination schedule. **E)** Distribution of neutralization levels one year after boosters 2 to 5 after annual half dose boosters. **F)** Comparison of neutralization level distributions during yearly measurements for annual full dose and biannual half-dose vaccination. **C–F)** Vertical lines: median neutralization levels. **C–F)** Blue and green bars: seniors. Red bars: healthcare workers.

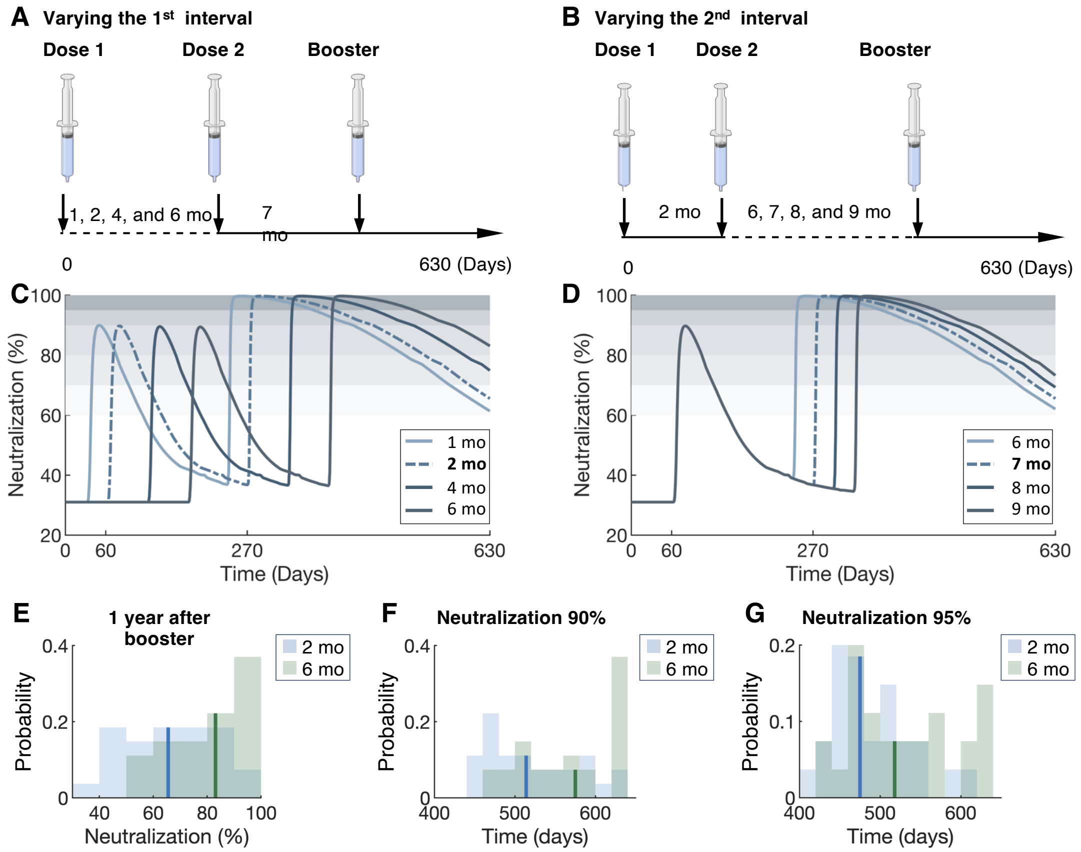


**Supplementary Figure 8. Predicted neutralization dynamics with varying dose intervals. A)** Diagram of a vaccination schedule with varying intervals between the first two doses (1, 2, 4, and 6 months). **B)** Diagram of a vaccination schedule with varying intervals between the second dose and the booster dose (6 to 9 months). **C)** Estimated median neutralization levels for seniors based on the intervals described in **A**. **D)** Estimated median neutralization levels for seniors based on the intervals described in **B**. Comparison between the current vaccination schedule and the regimen with a 6-month interval between the two priming doses for **E)** neutralization distributions on day 630, **F)** duration above the 90% threshold, and **G)** duration above the 95% threshold, respectively. **C–D)** Dashed lines: neutralization dynamics based on current vaccination guidelines. Shaded regions from light to dark gray: neutralization levels of 60-70%, 70-80%, 80-90%, 90-95%, and above 95%, respectively. **E–G)** Blue bars: current regimen. Green bars: regimen with an extended 6-month priming dose interval. Vertical lines: corresponding median values.
